# Prenatal alcohol exposure and the development of multiple risk behaviours in adolescence: A birth cohort study

**DOI:** 10.64898/2026.01.29.26345107

**Authors:** James T. I. Parsonage, Laura Tinner, David Troy, Caroline M. Taylor, Cheryl McQuire

## Abstract

**Background:** The UK has the fourth highest estimated prevalence globally of maternal alcohol consumption during pregnancy (41%). It is therefore important to understand the long-term impacts of prenatal alcohol exposure (PAE) by examining its impact on the development of adolescent multiple risk behaviours (MRBs) which may increase morbidity and premature mortality across the life-course.

**Methods:** Using Avon Longitudinal Study of Parents and Children (ALSPAC) cohort data with multiple imputation (n=6,752), we examined the impacts of ‘infrequent’, ‘frequent’, and ‘binge’ PAE groups on the development of seven MRBs at 16 years old, encompassing substance misuse, risky sexual behaviour and antisocial behaviour. Data were analysed using multiple regression, using q-statistics to adjust for multiple comparisons.

**Results:** Adolescents with ‘infrequent’ and ‘frequent’ PAE were more likely to develop hazardous alcohol use at 16 years old compared to those without PAE, with the strongest association being for the ‘frequent’ group (adjusted odds ratio (aOR) 1.45 [1.19-1.76], p<0.001, q-value=0.005). Adolescents exposed to binge drinking prenatally had an increased risk of engaging in underage sexual intercourse (aOR 1.34 [1.09-1.64], p=0.005, q-value=0.044). Binge drinking predicted a higher total MRB score (Coefficient = (+0.21 [+0.08 to +0.33], p=0.001, q-value=0.017).

**Conclusions:** This study supports the UK Chief Medical Officer’s Low Risk Drinking Guidelines that the safest approach if pregnant, or if there is a possibility of becoming pregnant, is to avoid drinking alcohol, with the more alcohol consumed during pregnancy the greater the risks of long-term harm to the baby. Given the findings that PAE may increase the risk of adolescent hazardous alcohol use and risky sexual behaviour, this study highlights the need for further research to understand the intergenerational effects of PAE.

## INTRODUCTION

The UK Chief Medical Officer’s (CMO) guidelines advise that the safest approach to mitigate the short and long-term risks of alcohol consumption during pregnancy is to avoid alcohol altogether (Public Health England, 2020). The prevalence of alcohol consumption during pregnancy in the UK has been estimated to be about 41%, one of the highest estimated percentages in Europe, which has a regional prevalence of fetal alcohol spectrum disorders (FASD) about 2.6 times higher than the global average (Popova et al., 2017). The prevalence of FASD has been estimated in UK school children at 1.8% and, when including ‘probable’ cases, was 3.6% (McCarthy et al., 2021).

Prenatal alcohol exposure (PAE) is a key factor in the diagnostic criteria for FASD and those ‘at risk of neurodevelopmental disorder or FASD’ (Scottish Intercollegiate Guidelines Network [hereafter SIGN], 2019). The neurodevelopmental disorder of FASD results in lifelong impairment and may present as problems with attention and cognition, social impairment, and difficulties regulating behaviour. For those with FASD, adolescence represents a crucial period where neurodevelopmental impairments meet growing societal pressures in the transition to adulthood, increasing the need for high intensity support (McLachlan et al., 2020). Exposure to and uptake of risk-taking behaviours may begin in adolescence as individuals develop greater autonomy over life choices affecting their health (Campbell et al., 2020).

The long-term impact of PAE on children in adolescence and beyond may be examined by using multiple risk behaviours (MRBs) as outcomes. MRBs develop or increase throughout adolescence and can prevail throughout life, leading to future morbidity and premature mortality (Wright et al., 2018). The risk to an individual’s health accumulates based on the number of distinct MRBs performed rather than pattern of different behaviours shown (Wright et al., 2020). As such, MRBs can be considered both as a total number of distinct risk behaviours, as well as individual behaviours that meet a threshold considered hazardous to an individual’s health.

The aetiology and occurrence of risk behaviours has been well explored in the literature on FASD. Individuals with FASD may struggle with executive functioning resulting in difficulties with socialisation, impulse control, and cause and effect thinking, and some may express behaviours including aggressiveness, criminal activity and sexual inappropriateness (Rasmussen et al., 2008). A recent meta-analysis found that conduct disorders, alcohol and drug dependence, and disturbances of activity or attention were extremely common in those with FASD (Popova et al., 2016). The Raine study found that four or more Australian standard drinks of alcohol per week (100ml of wine or equivalent) during pregnancy led to an increased risk of adolescent harmful alcohol use, compared to no PAE (Duko et al., 2020). The Mater University Study of Pregnancy found that three or more glasses a few times per month during pregnancy was correlated with alcohol use disorders in adolescence (Alati et al., 2006). Individuals with FASD also experience many secondary difficulties throughout their lives such as meeting independent living needs (Olsen and Sparrow, 2021). It is essential that strengths-based approaches and interventions are developed to improve the quality of life of those with the condition and the difficulties associated with it. This includes removing system-level barriers experienced by many families, carers and individuals with FASD (Petrenko et al., 2014). However, the need to understand the impacts of PAE remains, to target and improve services and approaches which support the health of those with FASD or PAE.

This study addresses this need by bringing the adolescent impacts of PAE into sharper focus and is the first to quantify the impacts of PAE with respect to a broad range of MRBs. Using Avon Longitudinal Study of Parents and Children (ALSPAC) birth cohort data, this observational study aims to examine seven MRB outcomes for adolescents with PAE. Our primary aim was to examine the impact of different patterns of PAE, including binge drinking, on the total number of MRBs, encompassing substance use, antisocial behaviour and risky sexual behaviour, displayed by age 16 years. Our secondary aim was to look at how different patterns of PAE impact the development of each individual risk behaviour.

## MATERIALS AND METHODS

### Overview of the data source and study design

Pregnant women resident in Avon, UK with expected dates of delivery between 1st April 1991 and 31st December 1992 were invited to take part in the study with the initial number of pregnancies enrolled being 14,541. However, the total sample size for analyses using any data collected after the age of seven is 15,447 pregnancies (15,658 foetuses) due to subsequent recruitment (Boyd et al., 2013; Fraser et al., 2013; Northstone et al., 2023). Please note that the study website contains details of all the data that is available through a fully searchable data dictionary and variable search tool (University of Bristol, 2025).

From an initial sample of 15,582 consented birth records, we excluded records of: a) participants from phase IV (children enrolled after the age of 18), b) children who died before one year of age, and c) mothers and partners assigned to the ‘armed forces’ social class, due to sparse data. 14,603 linked records of mothers, partners and their children remained eligible as shown in Figure 1. Maternal, paternal and child data were gathered via questionnaires and clinic assessments.

**Figure 1.**
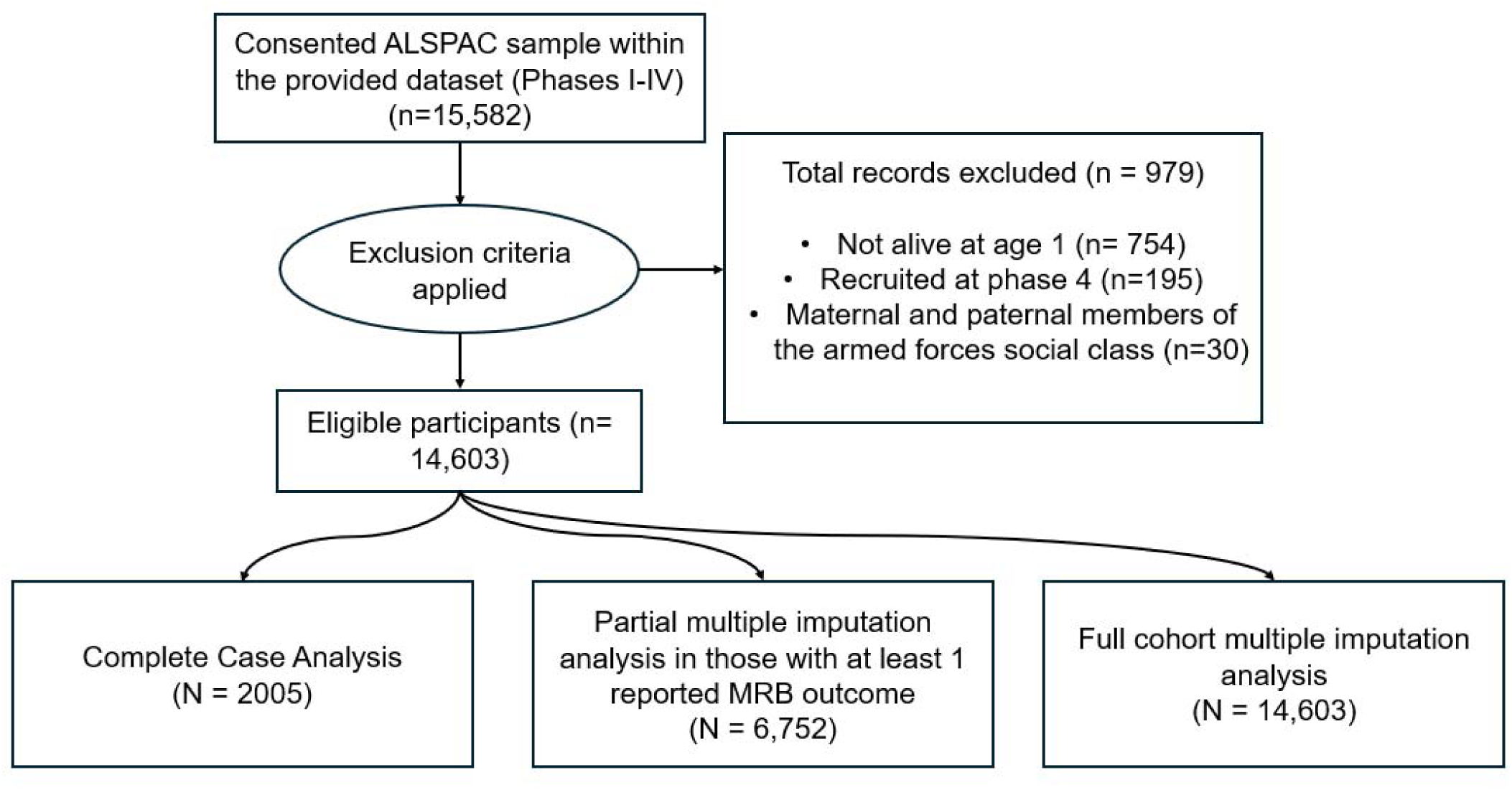
Participant flow diagram.

We first analysed data using a complete-case strategy, where only those with available data for all exposure, confounder and outcome variables were included. This resulted in a considerably attenuated sample size of 2,005 participants. To increase precision, explain missingness and address the notable bias in prenatal binge drinking exposure, maternal and paternal covariates and MRB outcomes when conditioning on being a complete case (shown in Supplementary Table 2), we performed multiple imputation under the assumption that the data were missing at random. We performed two imputation strategies, a partial imputation where individuals had at least one complete MRB outcome (n=6,752) and a full imputation for all eligible records in the ALSPAC cohort (n=14,603). There was substantial missingness found in the outcome MRBs, ranging between 64.7% and 69.0% of records in the full imputation. Given concerns around the degree of outcome missingness and therefore amount of imputed data in the full imputation, the partial multiple imputation analysis is considered the primary analysis. Results for the complete case analysis are provided in Supplementary Tables 3 and 4. The full ALSPAC imputation results are shown in Supplementary Tables 7 and 8. Figure 2 shows the substantive model and missingness model variables which are part of the complete case strategy and subsequent multiple imputation analyses.

**Figure 2.**
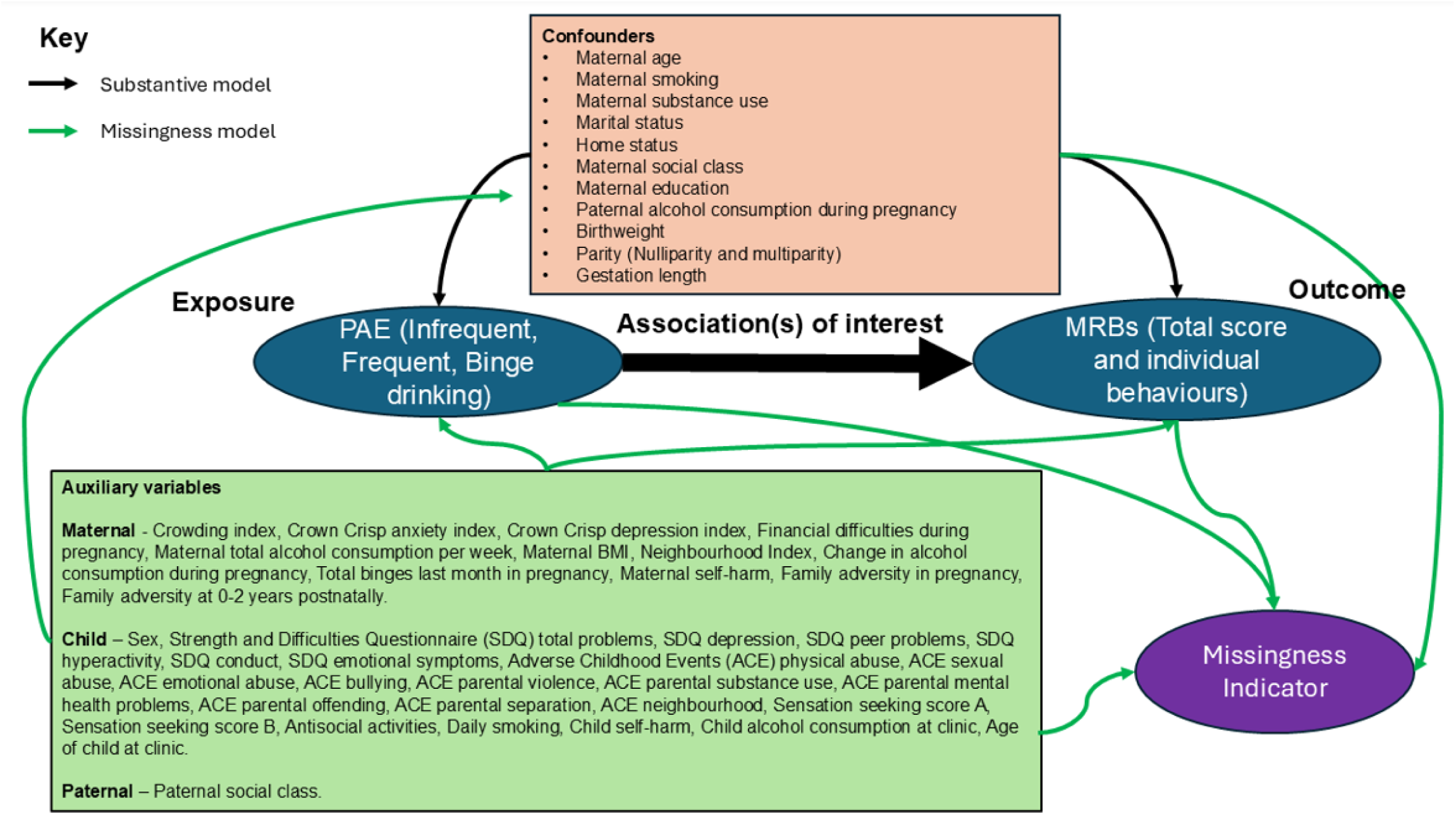
Included substantive model and missingness model variables.

### Ethical approval

The ALSPAC Executive Committee approved the study proposal on 29^th^ April 2024 (Proposal identifier: B4600). Ethical approval for the study was obtained from the ALSPAC Ethics and Law Committee and the Local Research Ethics Committees. Informed consent for the use of data collected via questionnaires and clinics was obtained from participants following the recommendations of the ALSPAC Ethics and Law Committee at the time.

### Exposure variables

Table 1 shows exposure definitions based on the timing, amount, and frequency of PAE, measured by a self-reported maternal questionnaire at 18 weeks gestation. Two exposure types were examined in this study. The first was increasing frequency of PAE grouped as: i) No PAE reported, ii) infrequent PAE (<1 glass of wine per week or equivalent measure), iii) frequent PAE (≥1 glass of wine per week or equivalent measure). The second was prenatal binge drinking exposure grouped as: i) No binge drinking reported, ii) Binge drinking (≥2 pints of beer or equivalent measure on one or more occasion).

**Table 1.**
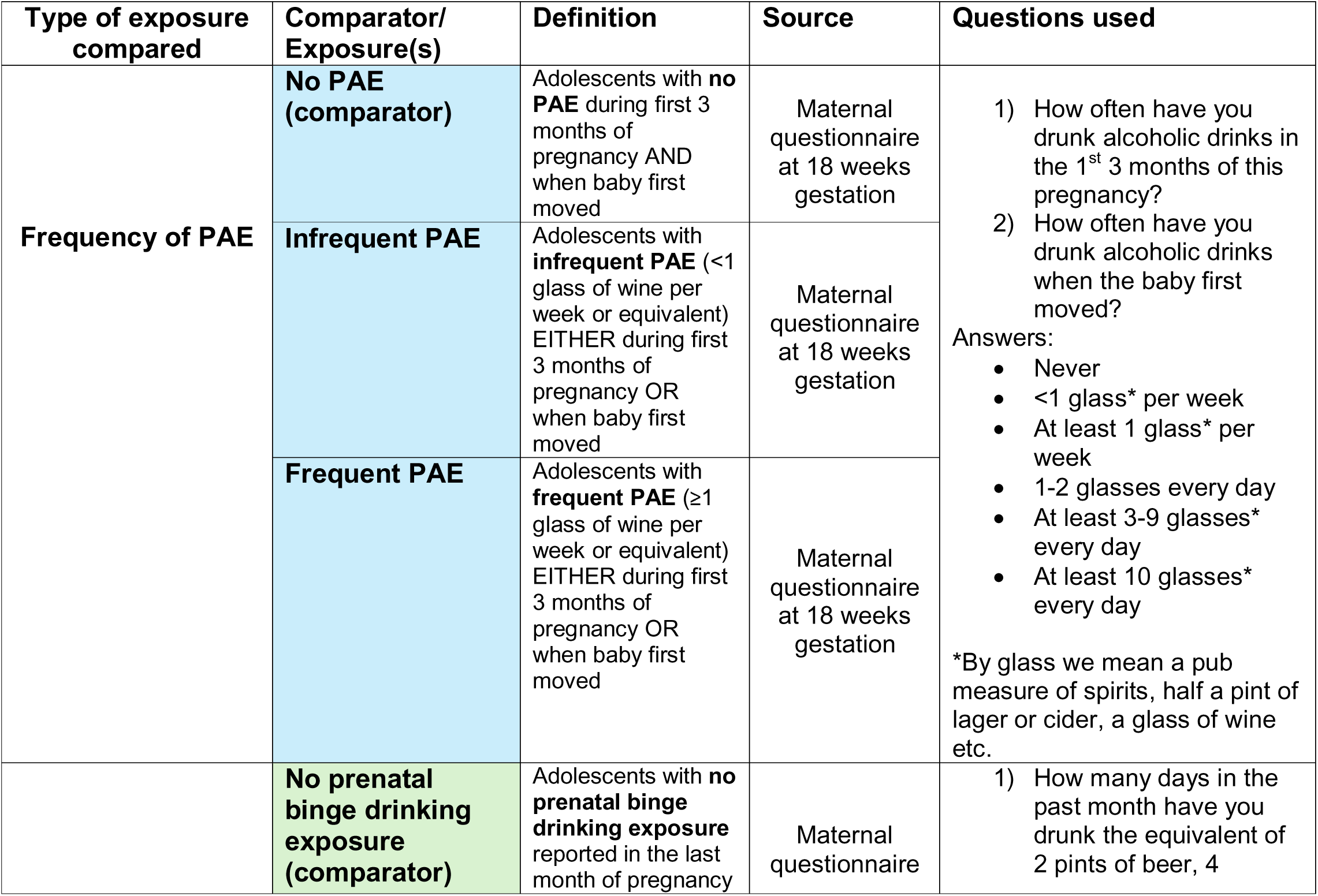
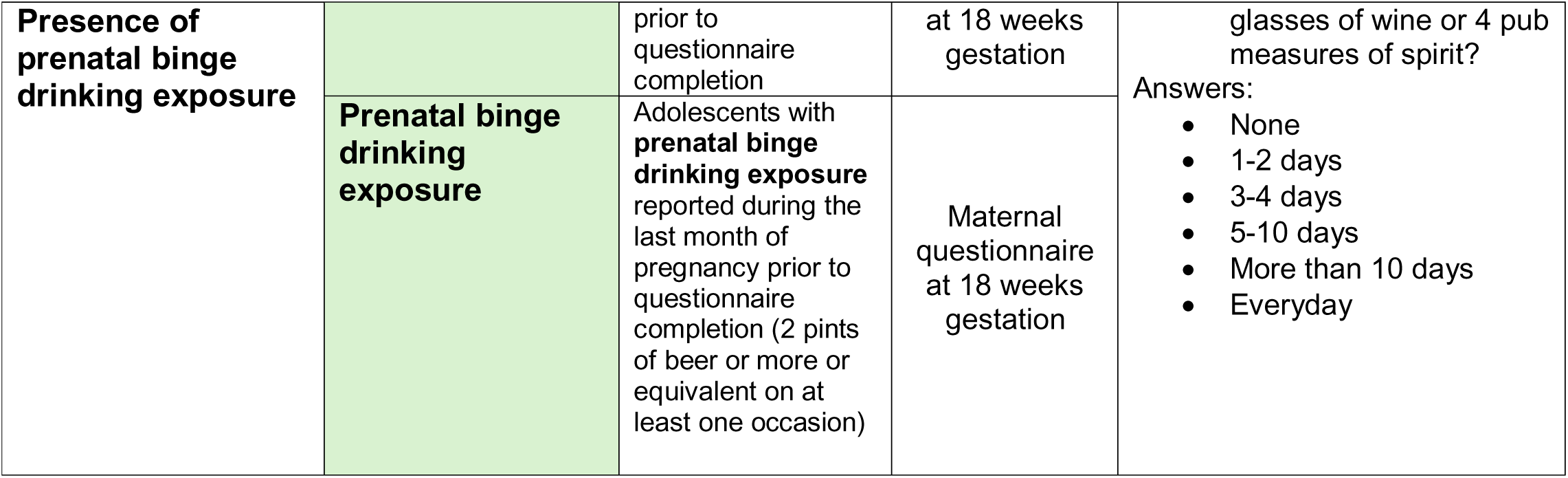
Prenatal alcohol exposure and comparator group definitions.

### Outcome variables

Outcomes were 7 MRBs encompassing substance use, antisocial, and risky sexual behaviour. Hazardous alcohol use, smoking, cannabis use, and drug use (excluding cannabis) outcomes were derived from a child self-reported questionnaire at age 16. Antisocial behaviour, underage sexual intercourse and unprotected sexual intercourse outcomes were derived from questions asked at clinic visits at age 15.5 years. Hazardous alcohol use questions collectively form the AUDIT score measure, a validated measure of problem drinking in adolescent populations (Rumph et al., 2013). All other outcomes use standardised MRB definitions, described in prior scientific literature (Wright et al., 2020). The definitions of each outcome are described in Table 2.

**Table 2.** ALSPAC definitions of Multiple Risk Behaviours.

| **Multiple Risk Behaviour** | **Definition** |
| --- | --- |
| **Hazardous alcohol use** | In the past year had scored 8 or more out of 40 on the Alcohol Use Disorders Identification Test (AUDIT) indicating hazardous alcohol consumption. |
| **Smoking** | Has ever smoked and is regularly smoking by currently smoking at least one cigarette per week. |
| **Cannabis use** | Those who reported using cannabis “sometimes but less often than once a week” or more regularly. |
| **Drug use (excluding cannabis)** | In the year since their 15th birthday, young person had either been a regular user (i.e. used five or more times) of one or more illicit drugs (excluding cannabis) including amphetamines, ecstasy, LSD, cocaine, ketamine or inhalants including aerosols, gas, solvents and poppers. |
| **Engagement in unsafe sex** | Penetrative sex without the use of contraception on the last occasion they had had sex in the past year. |
| **Underage sexual intercourse** | Young person has engaged in sexual intercourse in the last year, and they were aged less than 16 years old. |
| **Antisocial behaviour** | Young person reported that at least once in the past year they had undertaken at least one of the following 7 offences:-carried a weapon; physically hurt someone on purpose; stolen something; sold illicit substances to another person; damaged property belonging to someone else either by using graffiti, setting fire to it or destroying or damaging it in another fashion; subjected someone to verbal or physical racial abuse; or been rude/rowdy in a public place, |

### Covariates and confounding

Drawing on the literature, we developed a directed acyclic graph (DAG) using DAGitty software to depict hypothesised causal and confounding pathways and inform covariate selection to minimise bias in our substantive models (Figure 3) (Textor et al., 2016; McQuire et al., 2019). Covariates used for model adjustment were maternal age, parity, birthweight, maternal social class, marital status, home ownership status, highest maternal educational qualification, maternal smoking, maternal cannabis use, maternal drug use (excluding cannabis), and paternal alcohol use during pregnancy. Likelihood ratio tests, for each MRB, were used to ascertain that the adjusted model was a better fit for the observed data than an unadjusted model.

**Figure 3.**
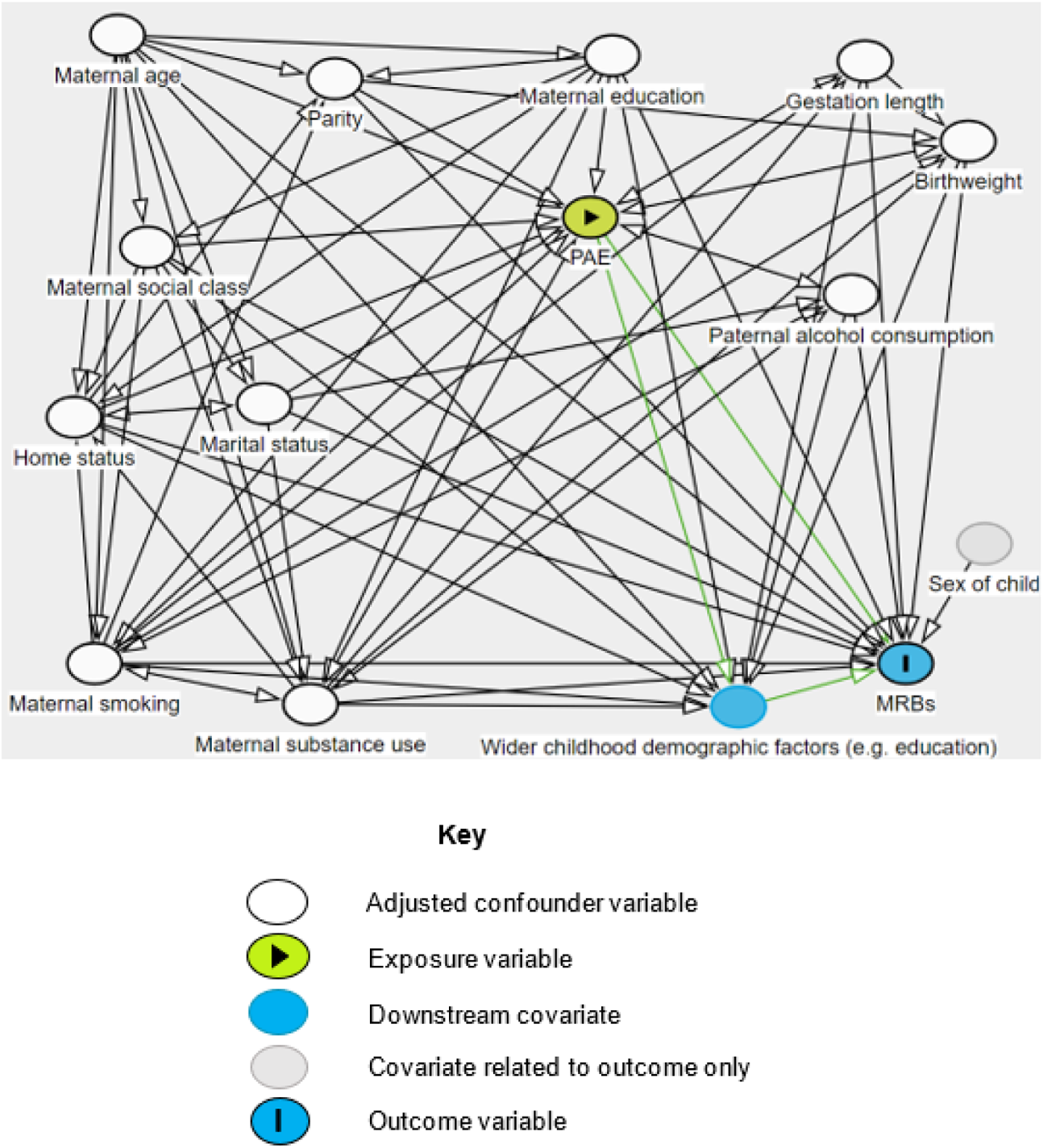
Directed acyclic graph of confounding covariates between PAE and MRBs.

### Descriptive statistics

Tables 3 and 4 show pooled descriptive statistics from the multiple imputation. We calculated descriptive statistics for the complete cases using the ‘arsenal’ R package (Heinzen, 2016). Descriptive statistics for complete cases and non-complete cases are presented in Supplementary Tables 1 and 2 respectively.

**Table 3.** Multiple imputation sample characteristics table for the component-based MRB analysis of individuals with at least one complete MRB variable (N = 6,752)

|  | Frequency of prenatal alcohol exposure (PAE) |  |  |  |  | Prenatal binge drinking exposure groups |  |  |  |
| --- | --- | --- | --- | --- | --- | --- | --- | --- | --- |
| Maternal, child or partner characteristics | No PAE<br>(N=2,268) | Infrequent PAE<br>(N=3,552) | Frequent PAE<br>(N=932) | Total<br>(N=6,752) | p-value<sup>a</sup> | No binge drinking<br>(N=5,769) | Binge drinking<br>(N=983) | Total<br>(N=6,752) | p-value<sup>a</sup> |
| **Maternal age at delivery (years)** |  |  |  |  |  |  |  |  |  |
| ≤19 years | 59<br>(2.6%) | 51<br>(1.4%) | 9<br>(0.9%) | 119<br>(1.8%) | **<0.001** | 99<br>(1.7%) | 20<br>(2.0%) | 119<br>(1.8%) | **0.20** |
| >19 years and ≤ 34 years | 2,004<br>(88.4%) | 3,045<br>(85.7%) | 736<br>(78.9%) | 5,785<br>(85.7%) |  | 4,960<br>(86.0%) | 824<br>(83.8%) | 5,785<br>(85.7%) |  |
| ≥35 years | 204<br>(9.0%) | 457<br>(12.9%) | 188<br>(20.2%) | 849<br>(12.6%) |  | 709<br>(12.3%) | 139<br>(14.1%) | 849<br>(12.6%) |  |
| **Any smoking during pregnancy** |  |  |  |  |  |  |  |  |  |
| No | 1,925<br>(84.9%) | 2,943<br>(82.8%) | 711<br>(76.3%) | 5,579<br>(82.6%) | **<0.001** | 4,917<br>(85.2%) | 662<br>(67.3%) | 5,579<br>(82.6%) | **<0.001** |
| Yes | 343<br>(15.1%) | 609<br>(17.2%) | 221<br>(23.7%) | 1,173<br>(17.4%) |  | 852<br>(14.8%) | 321<br>(32.7%) | 1,173<br>(17.4%) |  |
| **Maternal cannabis use during pregnancy** |  |  |  |  |  |  |  |  |  |
| No | 2,246<br>(99.0%) | 3,476<br>(97.8%) | 881<br>(94.5%) | 6,603<br>(97.8%) | **<0.001** | 5,665<br>(98.2%) | 938<br>(95.4%) | 6,603<br>(97.8%) | **<0.001** |
| Yes | 22<br>(1.0%) | 76<br>(2.2%) | 51<br>(5.5%) | 149<br>(2.2%) |  | 104<br>(1.8%) | 46<br>(4.6%) | 149<br>(2.2%) |  |
| **Maternal drug use (excluding cannabis) during pregnancy** |  |  |  |  |  |  |  |  |  |
| No | 2260<br>(99.7%) | 3,543<br>(99.7%) | 925<br>(99.2%) | 6,729<br>(99.7%) | **0.07** | 5,754<br>(99.7%) | 975<br>(99.1%) | 6,729<br>(99.7%) | **0.007** |
| Yes | 7<br>(0.3%) | 9<br>(0.3%) | 7<br>(0.8%) | 23<br>(0.3%) |  | 15<br>(0.3%) | 8<br>(0.9%) | 23<br>(0.3%) |  |
| **Parity** |  |  |  |  |  |  |  |  |  |
| Nulliparous | 1,182<br>(52.1%) | 1,634<br>(46.0%) | 439<br>(47.1%) | 3,255<br>(48.2%) | **<0.001** | 2,813<br>(48.8%) | 442<br>(45.0%) | 3,255<br>(48.2%) | **<0.001** |
| Primiparous | 757<br>(33.4%) | 1,304<br>(36.7%) | 306<br>(32.9%) | 2,367<br>(35.1%) |  | 2,051<br>(35.6%) | 315<br>(32.1%) | 2,367<br>(35.1%) |  |
| Multiparous | 329<br>(14.5%) | 614<br>(17.3%) | 186<br>(20.0%) | 1,129<br>(16.7%) |  | 904<br>(15.7%) | 226<br>(23.0%) | 1,129<br>(16.7%) |  |
| **Maternal social class** |  |  |  |  |  |  |  |  |  |
| I to III (Non-manual) | 1,812<br>(79.9%) | 2,966<br>(83.5%) | 807<br>(86.5%) | 5,585<br>(82.7%) | **<0.001** | 4,817<br>(83.5%) | 768<br>(78.1%) | 5,585<br>(82.7%) | **<0.001** |
| III (Manual) to V | 455<br>(20.1%) | 586<br>(16.5%) | 126<br>(13.5%) | 1,167<br>(17.3%) |  | 952<br>(16.5%) | 216<br>(21.9%) | 1,167<br>(17.3%) |  |
| **Mother's highest educational qualification** |  |  |  |  |  |  |  |  |  |
| A-level or above | 849<br>(37.4%) | 1,679<br>(47.3%) | 498<br>(53.4%) | 3,026<br>(44.8%) | **<0.001** | 2,668<br>(46.2%) | 359<br>(36.5%) | 3,026<br>(44.8%) | **<0.001** |
| Lower than A-level | 1,419<br>(62.6%) | 1,873<br>(52.7%) | 434<br>(46.6%) | 3,726<br>(55.2%) |  | 3,101<br>(53.8%) | 625<br>(63.5%) | 3,726<br>(55.2%) |  |
| **Home ownership status** |  |  |  |  | **0.01** |  |  |  | **<0.001** |
| Own home | 1,851<br>(81.6%) | 3,004<br>(84.6%) | 783<br>(84.0%) | 5,688<br>(83.5%) |  | 4,880<br>(84.6%) | 758<br>(77.1%) | 5,688<br>(83.5%) |  |
| No owned home or mortgage | 417<br>(18.4%) | 548<br>(15.4%) | 149<br>(16.0%) | 1,114<br>(16.5%) |  | 888<br>(15.4%) | 226<br>(22.9%) | 1,114<br>(16.5%) |  |
| **Marital status** |  |  |  |  |  |  |  |  |  |
| Married | 1,865<br>(82.3%) | 2,928<br>(82.4%) | 706<br>(75.7%) | 5,499<br>(81.4%) | <0.001 | 4,803<br>(83.3%) | 697<br>(70.8%) | 5,499<br>(81.4%) | <0.001 |
| Not married | 402<br>(17.7%) | 624<br>(17.6%) | 227<br>(24.3%) | 1,253<br>(18.6%) |  | 966<br>(16.7%) | 287<br>(29.2%) | 1,253<br>(18.6%) |  |
| **Sex of child** |  |  |  |  |  |  |  |  |  |
| Female | 1,250<br>(55.1%) | 1,958<br>(55.1%) | 504<br>(54.1%) | 3,713<br>(55.0%) | 0.83 | 3,167<br>(54.9%) | 546<br>(55.5%) | 3,713<br>(55.0%) | 0.74 |
| Male | 1,017<br>(44.9%) | 1,594<br>(44.9%) | 428<br>(45.9%) | 3,039<br>(45.0%) |  | 2,602<br>(45.1%) | 437<br>(44.5%) | 3,039<br>(45.0%) |  |
| **Child birthweight** |  |  |  |  |  |  |  |  |  |
| ≥2,500 grams | 2,138<br>(94.3%) | 3,393<br>(95.5%) | 882<br>(94.6%) | 6,414<br>(95.0%) | 0.10 | 5,487<br>(95.1%) | 927<br>(94.2%) | 6,414<br>(95.0%) | 0.27 |
| <2,500 grams | 129<br>(5.7%) | 159<br>(4.5%) | 50<br>(5.4%) | 338<br>(0.5%) |  | 282<br>(4.9%) | 57<br>(5.8%) | 338<br>(0.5%) |  |
| **Length of pregnancy (gestational length)** |  |  |  |  |  |  |  |  |  |
| Extreme pre-term | * | * | * | * | 0.16 | * | * | * | 0.29 |
| Pre-term | * | * | * | * |  | * | * | * |  |
| Term | 2,130<br>(93.9%) | 3,374<br>(95.0%) | 881<br>(94.5%) | 6,385<br>(94.6%) |  | 5,460<br>(94.7%) | 925<br>(94.0%) | 6,385<br>(94.6%) |  |
| **Partner's alcohol consumption during pregnancy** |  |  |  |  |  |  |  |  |  |
| Never | 181<br>(8.0%) | 71<br>(2.0%) | 7<br>(0.8%) | 259<br>(3.8%) | <0.001 | 248<br>(4.8%) | 12<br>(1.2%) | 259<br>(3.8%) | <0.001 |
| Infrequent | 722<br>(31.9%) | 766<br>(21.6%) | 58<br>(6.2%) | 1,546<br>(22.9%) |  | 1,409<br>(24.4%) | 137<br>(13.9%) | 1,546<br>(22.9%) |  |
| Frequent | 1,364<br>(60.2%) | 2,715<br>(76.4%) | 867<br>(93.0%) | 4,946<br>(73.3%) |  | 4,111<br>(71.3%) | 835<br>(84.9%) | 4,946<br>(73.3%) |  |
| **MRB - Hazardous alcohol use** |  |  |  |  |  |  |  |  |  |
| No | 1,538<br>(67.8%) | 2,195<br>(61.8%) | 499<br>(53.6%) | 4,232<br>(62.7%) | <0.001 | 3,687<br>(63.9%) | 545<br>(55.4%) | 4,232<br>(62.7%) | <0.001 |
| Yes | 730<br>(32.2%) | 1,357<br>(38.2%) | 433<br>(46.4%) | 2,520<br>(37.3%) |  | 2,082<br>(36.1%) | 438<br>(44.6%) | 2,520<br>(37.3%) |  |
| **MRB - Drug use (excluding cannabis)** |  |  |  |  |  |  |  |  |  |
| No | 2,136<br>(94.2%) | 3,330<br>(93.7%) | 853<br>(91.5%) | 6,319<br>(93.6%) | 0.02 | 5,425<br>(94.0%) | 894<br>(90.9%) | 6,319<br>(93.6%) | <0.001 |
| Yes | 132<br>(5.8%) | 222<br>(6.3%) | 79<br>(8.5%) | 433<br>(6.4%) |  | 344<br>(6.0%) | 89<br>(9.1%) | 433<br>(6.4%) |  |
| **MRB - Cannabis use** |  |  |  |  |  |  |  |  |  |
| No | 2,077<br>(91.6%) | 3,159<br>(88.9%) | 809<br>(86.8%) | 6,045<br>(89.5%) | <0.001 | 5,192<br>(90.0%) | 853<br>(86.7%) | 6,045<br>(89.5%) | 0.002 |
| Yes | 190<br>(8.4%) | 393<br>(11.1%) | 123<br>(13.2%) | 707<br>(10.5%) |  | 577<br>(10.0%) | 130<br>(13.3%) | 707<br>(10.5%) |  |
| **MRB - Regular smoking** |  |  |  |  | 0.10 |  |  |  | <0.001 |
| No | 1,965<br>(86.6%) | 3,081<br>(86.8%) | 784<br>(84.1%) | 5,830<br>(86.3%) |  | 5,040<br>(87.4%) | 791<br>(80.4%) | 5,830<br>(86.3%) |  |
| Yes | 303<br>(13.4%) | 470<br>(13.2%) | 148<br>(15.9%) | 922<br>(13.7%) |  | 729<br>(12.6%) | 193<br>(19.6%) | 922<br>(13.7%) |  |
| **MRB -**<br>**Antisocial**<br>**behaviour** |  |  |  |  |  |  |  |  |  |
| No | 1,263<br>(55.7%) | 1,874<br>(52.8%) | 487<br>(52.2%) | 3,624<br>(53.7%) | **0.06** | 3,153<br>(54.7%) | 472<br>(48.0%) | 3,624<br>(53.7%) | **<0.001** |
| Yes | 1,004<br>(44.3%) | 1,678<br>(47.2%) | 445<br>(47.8%) | 3,128<br>(46.3%) |  | 2,616<br>(45.3%) | 512<br>(52.0%) | 3,128<br>(46.3%) |  |
| **MRB - Unsafe**<br>**sex** |  |  |  |  |  |  |  |  |  |
| No | 2,222<br>(98.0%) | 3,488<br>(98.2%) | 919<br>(98.6%) | 6,629<br>(98.2%) | **0.56** | 5,671<br>(98.3%) | 958<br>(97.4%) | 6,629<br>(98.2%) | **0.07** |
| Yes | 45<br>(2.0%) | 64<br>(1.8%) | 13<br>(1.4%) | 122<br>(1.8%) |  | 98<br>(1.7%) | 25<br>(2.6%) | 122<br>(1.8%) |  |
| **MRB -**<br>**Underage sex** |  |  |  |  |  |  |  |  |  |
| No | 1,880<br>(82.9%) | 2932<br>(82.6%) | 766<br>(82.2%) | 5,578<br>(82.6%) | **0.88** | 4,828<br>(83.7%) | 751<br>(76.3%) | 5,578<br>(82.6%) | **<0.001** |
| Yes | 388<br>(17.1%) | 620<br>(17.4%) | 166<br>(17.8%) | 1,174<br>(17.4%) |  | 941<br>(16.3%) | 233<br>(23.7%) | 1,174<br>(17.4%) |  |
| **Abbreviations: PAE = prenatal alcohol exposure, MRB = multiple risk behaviour**<br><sup>a</sup> P-values calculated using chi-squared analyses for categorical variables.<br>The * sign indicates suppressed counts of <5 people, or counts where a suppressed number can be calculated e.g. from the percentage of another count.<br>Participant counts may not sum to the row total N nor the column header total N, as values are based on pooled multiple imputation estimates rounded to the nearest integer. Column header totals are also rounded imputed values with the exception of the sample total N = 6,752.<br>Percentages by covariate or MRB group may not total 100% due to percentage rounding to 1 decimal place. |  |  |  |  |  |  |  |  |  |

**Table 4.**
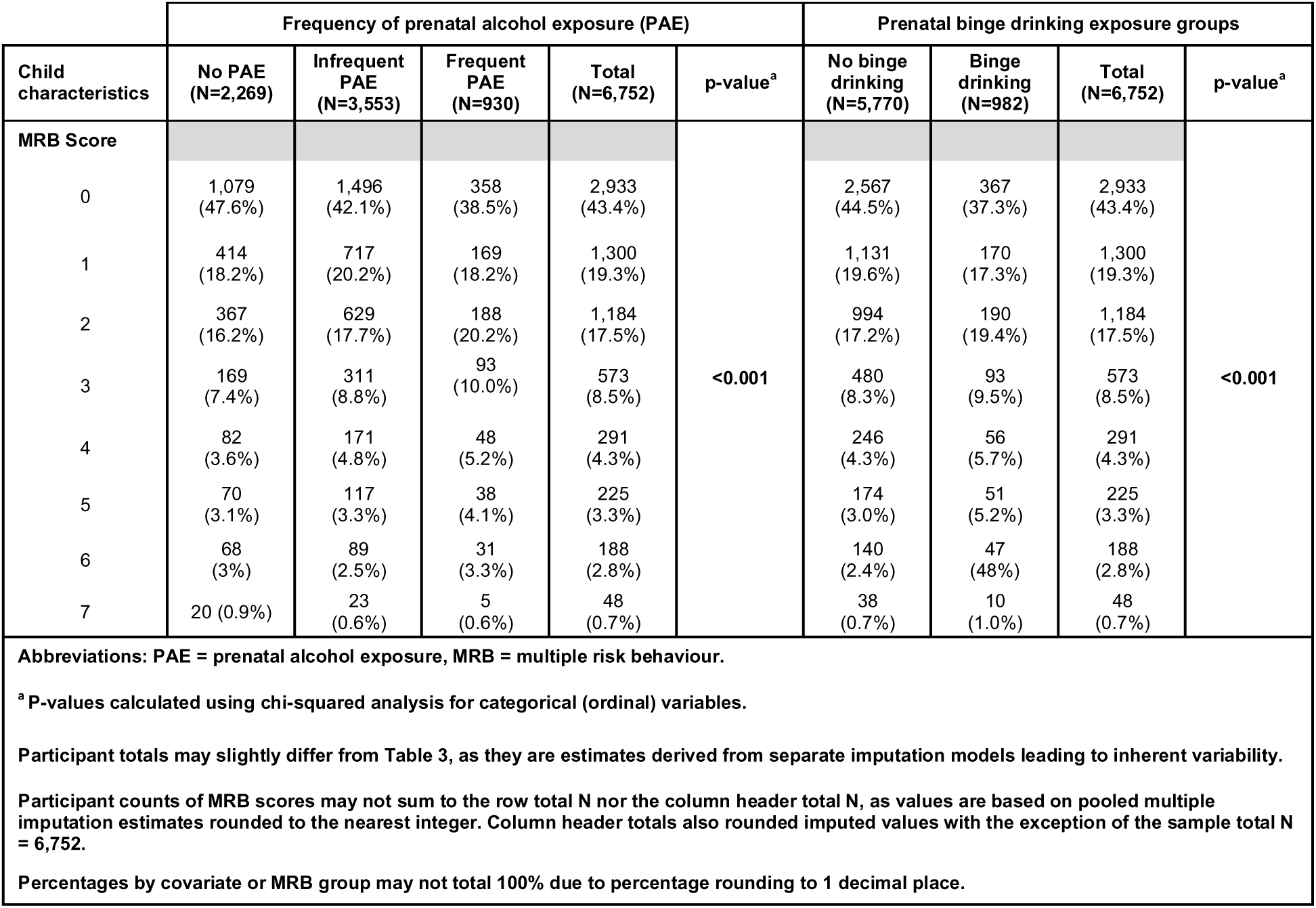
Descriptive statistics of passively imputed adolescent MRB scores by prenatal alcohol exposure groups for those with at least one complete MRB variable (N=6,752)

|  | Frequency of prenatal alcohol exposure (PAE) |  |  |  |  | Prenatal binge drinking exposure groups |  |  |  |
| --- | --- | --- | --- | --- | --- | --- | --- | --- | --- |
| Child characteristics | No PAE<br>(N=2,269) | Infrequent PAE<br>(N=3,553) | Frequent PAE<br>(N=930) | Total<br>(N=6,752) | p-value<sup>a</sup> | No binge drinking<br>(N=5,770) | Binge drinking<br>(N=982) | Total<br>(N=6,752) | p-value<sup>a</sup> |
| MRB Score |  |  |  |  |  |  |  |  |  |
| 0 | 1,079<br>(47.6%) | 1,496<br>(42.1%) | 358<br>(38.5%) | 2,933<br>(43.4%) | <0.001 | 2,567<br>(44.5%) | 367<br>(37.3%) | 2,933<br>(43.4%) | <0.001 |
| 1 | 414<br>(18.2%) | 717<br>(20.2%) | 169<br>(18.2%) | 1,300<br>(19.3%) |  | 1,131<br>(19.6%) | 170<br>(17.3%) | 1,300<br>(19.3%) |  |
| 2 | 367<br>(16.2%) | 629<br>(17.7%) | 188<br>(20.2%) | 1,184<br>(17.5%) |  | 994<br>(17.2%) | 190<br>(19.4%) | 1,184<br>(17.5%) |  |
| 3 | 169<br>(7.4%) | 311<br>(8.8%) | 93<br>(10.0%) | 573<br>(8.5%) |  | 480<br>(8.3%) | 93<br>(9.5%) | 573<br>(8.5%) |  |
| 4 | 82<br>(3.6%) | 171<br>(4.8%) | 48<br>(5.2%) | 291<br>(4.3%) |  | 246<br>(4.3%) | 56<br>(5.7%) | 291<br>(4.3%) |  |
| 5 | 70<br>(3.1%) | 117<br>(3.3%) | 38<br>(4.1%) | 225<br>(3.3%) |  | 174<br>(3.0%) | 51<br>(5.2%) | 225<br>(3.3%) |  |
| 6 | 68<br>(3%) | 89<br>(2.5%) | 31<br>(3.3%) | 188<br>(2.8%) |  | 140<br>(2.4%) | 47<br>(4.8%) | 188<br>(2.8%) |  |
| 7 | 20 (0.9%) | 23<br>(0.6%) | 5<br>(0.6%) | 48<br>(0.7%) |  | 38<br>(0.7%) | 10<br>(1.0%) | 48<br>(0.7%) |  |

Abbreviations: PAE = prenatal alcohol exposure, MRB = multiple risk behaviour.
<sup>a</sup> P-values calculated using chi-squared analysis for categorical (ordinal) variables.
Participant totals may slightly differ from Table 3, as they are estimates derived from separate imputation models leading to inherent variability.
Participant counts of MRB scores may not sum to the row total N nor the column header total N, as values are based on pooled multiple imputation estimates rounded to the nearest integer. Column header totals also rounded imputed values with the exception of the sample total N = 6,752.
Percentages by covariate or MRB group may not total 100% due to percentage rounding to 1 decimal place.

### Multiple imputation

Variables for multiple imputation models included hypothesised risk factors for PAE and MRBs, sociodemographic variables, and auxiliary variables (i.e. variables that are not included in the main analyses of interest, but which may improve the performance of missing data models) (see Figure 2) (Collins et al., 2001). Through systematically appraising the ALSPAC variable catalogue, a wide range of variables were selected which were considered to help explain missingness in substantive model variables. The ‘quickpred-pt2’ method was then used to assign variables to the imputation model with a correlation cut off (‘mincor’) of 0.2. The partial imputation presented here included all records with at least one MRB outcome (n=6,752) and therefore removes all individuals who had no MRB data.

Multiple imputation was performed using the MICE package in R software (van Buuren and Groothuis-Oudshoorn, 2011; R Core Team, 2023). For the MRB score analysis, 70 imputations were performed across 80 iterations for stability. For the individual MRB analysis, 50 imputations were performed across 50 iterations for stability. Rubins rules were applied to pool estimates across imputations for the multiple linear and logistic regression analyses. Further information about the imputation models, MICE methods, and diagnostics can be found in the supplementary material.

### Analysis

We analysed the imputed data using multiple regression to quantify the strength of the associations between PAE and the following outcomes at age 16 years: 1) the adjusted mean difference in MRB score in the PAE exposure groups compared with comparator groups (ranging from 0 to 7 MRBs); 2) the relative likelihood (adjusted odds ratio (aOR)) of participants displaying each component MRB between PAE exposure and comparator groups. We applied q-values to results to account for multiple comparisons (Storey, 2011).

## RESULTS

### Descriptive statistics

Within the multiple imputation sample (Table 3), 66.4% of mothers reported at least infrequent PAE. Mothers of children with increasing frequency of PAE were more likely to be older, to smoke, use cannabis, be unmarried, have higher educational qualifications, to be part of social classes I to III (Non-manual) and to have partners who drank more frequently during the pregnancy. Multiparity was more prevalent in mothers of children with frequent PAE, whereas nulliparity was more prevalent in mothers of children without PAE. The strongest association to increasing maternal alcohol consumption was their partner’s alcohol consumption.

A similar direction in associations was seen in mothers of children with binge drinking exposure compared with those without binge drinking exposure, with regards to maternal smoking, substance use and marital status. However higher percentages of mothers of children with binge drinking exposure were from social classes III-Manual to V (unskilled manual) and had a highest education qualification below A-level than mothers of children unexposed to binge drinking. There were no meaningful differences in maternal age at delivery and parity between mothers of children with binge drinking PAE compared with those who were unexposed to binge drinking.

### Multiple regression

#### Analysis 1: Adolescent MRB scores and PAE

There was no strong statistical evidence that children with ‘infrequent’ or ‘frequent’ PAE exhibited more MRBs at age 16 years than those without PAE (Table 5).

**Table 5.**
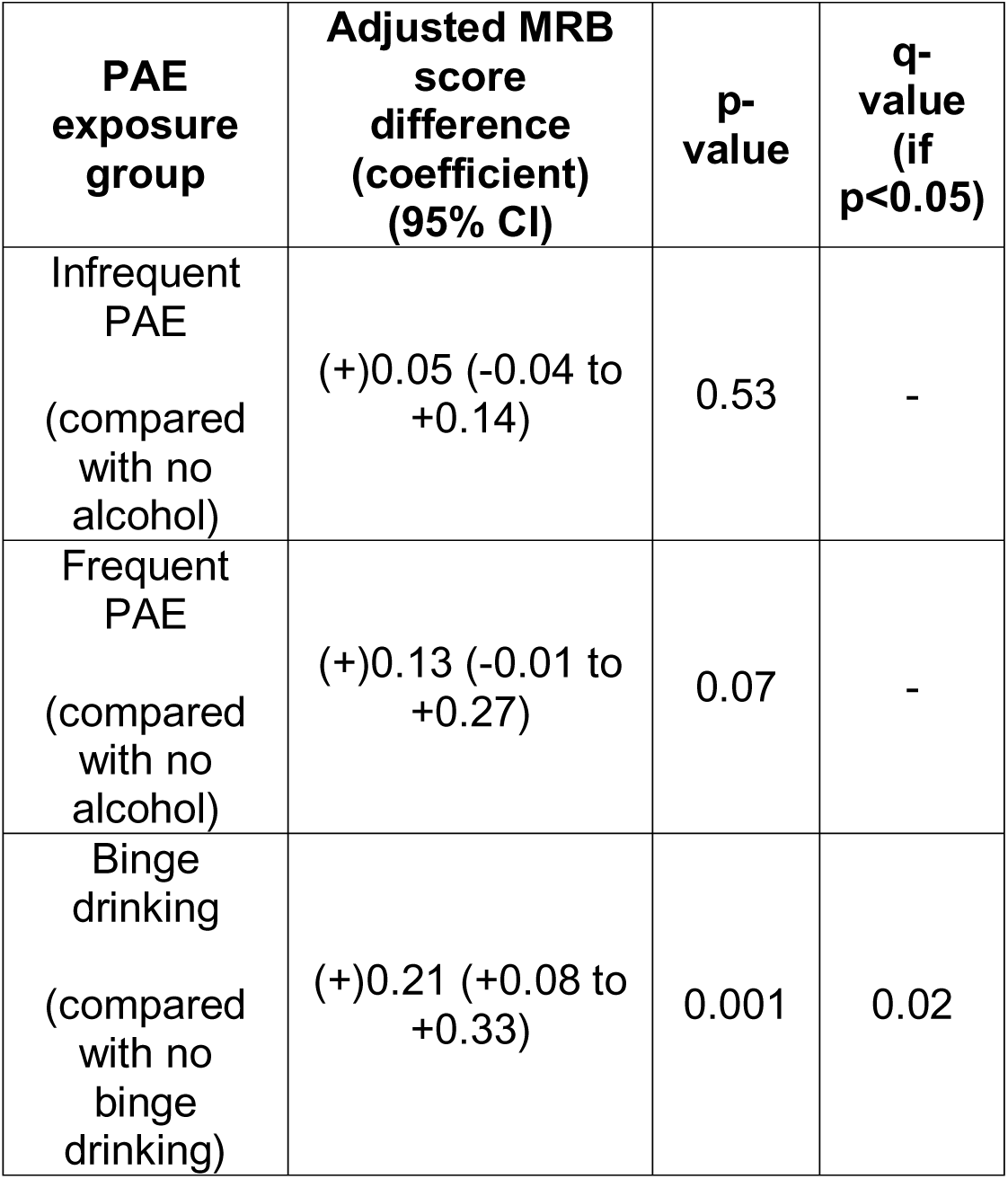
Multiple imputation results (N = 6,752) comparing adjusted MRB score between PAE exposure and comparator groups.

| **PAE exposure group** | **Adjusted MRB score difference (coefficient) (95% CI)** | **p-value** | **q-value (if p<0.05)** |
| --- | --- | --- | --- |
| Infrequent PAE<br>(compared with no alcohol) | (+)0.05 (-0.04 to +0.14) | 0.53 | - |
| Frequent PAE<br>(compared with no alcohol) | (+)0.13 (-0.01 to +0.27) | 0.07 | - |
| Binge drinking<br>(compared with no binge drinking) | (+)0.21 (+0.08 to +0.33) | 0.001 | 0.02 |

There was statistical evidence of a positive association between exposure to prenatal binge drinking (defined as two or more pints of beer or pub measure equivalent on at least 1 occasion) and MRBs in adolescents. Adolescents who were exposed to binge drinking prenatally had a small increase in adjusted mean MRB score ((+)0.21 (+0.08 to +0.33), p=0.001, q=0.017) compared to those unexposed to binge drinking.

#### Analysis 2: Individual risk behaviour development at adolescence and PAE

Table 6 shows the increased likelihood of hazardous alcohol use in the past year from age 16 years in adolescents with ‘frequent’ PAE, defined as ≥1 glass of wine per week or equivalent pub measure (aOR 1.45, 95% CI 1.19-1.76, p<0.001, q-value=0.005), compared with adolescents without PAE (Table 6). No evidence was found of an increased likelihood of developing other multiple risk behaviours at age 16 in either the ‘infrequent’ or ‘frequent’ PAE groups compared to adolescents without PAE.

**Table 6.**
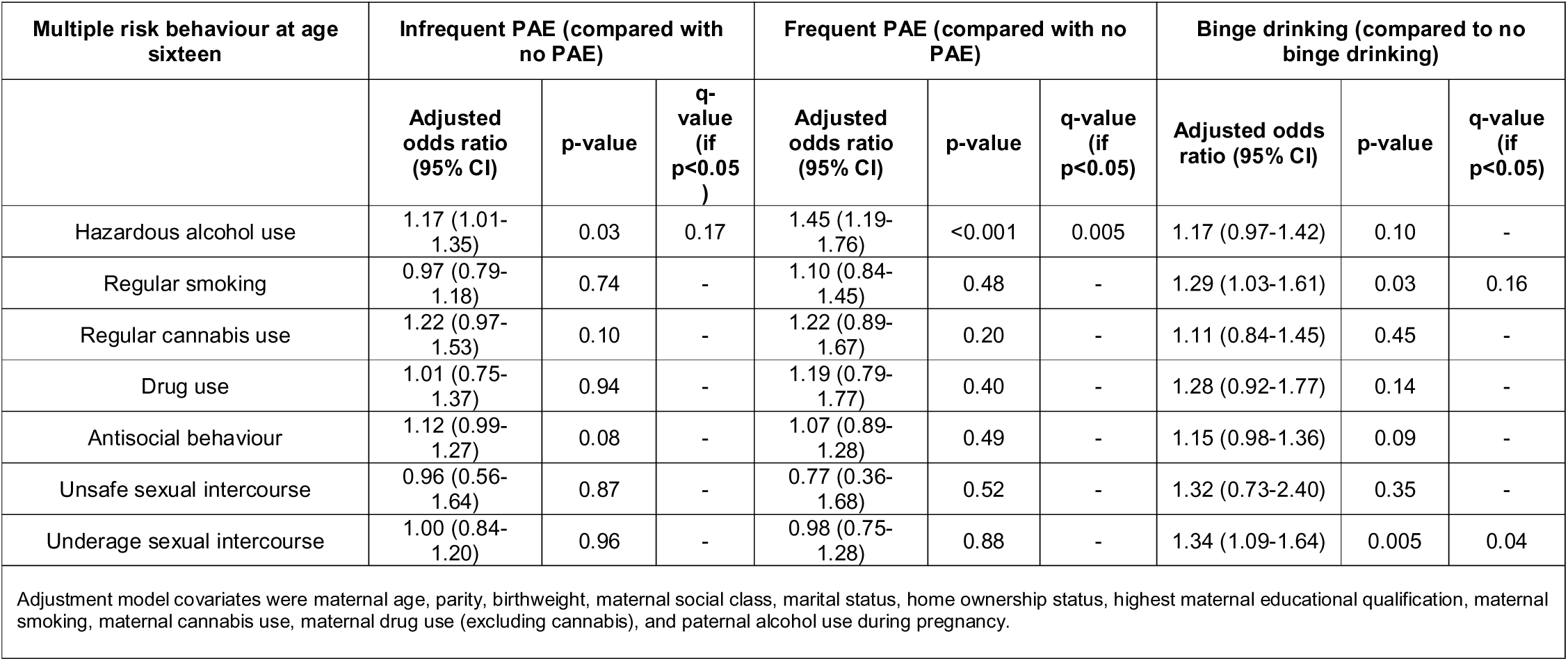
Multiple imputed (N = 6,752) aORs of MRBs in PAE exposure groups relative to comparator groups.

Adolescents exposed to binge drinking during pregnancy were more likely to engage in underage sexual intercourse by age 16 (aOR 1.34, 95% CI 1.09-1.64, p=0.005, q = 0.044) than adolescents unexposed to binge drinking during pregnancy. No q-value supported evidence was found that prenatal binge drinking exposure increased the likelihood of developing other MRBs at age 16 years.

## DISCUSSION

This study introduces important risky sexual behaviour outcomes, which were not included in a recent systematic review of ALSPAC studies investigating the link between PAE and childhood outcomes (Ogunjimi et al., 2025). It also provides different findings to the one study in the review investigating the association between PAE and problematic adolescent alcohol use which previously found that only in univariate analysis, maternal alcohol consumption was only related to adolescent alcohol consumption and adolescent alcohol problems (Kendler et al., 2013). However, differences may be explained by our use of a different outcome threshold, adolescent hazardous alcohol use (AUDIT score of 8 or more), and in our use of different ordinal groups by both amount, type and timing of prenatal alcohol exposure.

In our multivariate analysis, at a q-value level, the strongest relationship found was between exposure to frequent PAE (>=1 glass of wine per week or equivalent measure) and having higher odds of reported hazardous alcohol use at age 16. Binge-level PAE was associated with a small increase in overall number of risk behaviours developed and underage sexual intercourse in adolescence. As well as the increase in mean adolescent MRB scores that we observed in the binge drinking group in our study, the specific combination of types of MRB observed - hazardous alcohol use in the frequent drinking group and underage sexual intercourse in the group exposed to binge drinking prenatally – are of particular importance.

Concurrent hazardous alcohol use and underage sexual intercourse in adolescence may perpetuate intergenerational transmission of PAE and alcohol misuse.

It is also important to note that the sample prevalence of PAE estimated using multiple imputation in this study (66.4%) is likely reasonable when considering a modern UK prevalence of 41% may be considered conservative, with the upper bounds of modern estimates being up to 75% (Popova et al., 2017).

The exposure and comparator group descriptive differences are not unexpected. Increasing alcohol consumption is widely linked with smoking and cannabis use. Greater maternal age has also been associated with increased alcohol consumption in pregnancy (Meschke et al., 2013). In a US survey, those who were not married, were employed, and had higher educational attainment were more likely to report alcohol consumption during pregnancy (Gosdin et al., 2022). Binge drinking during pregnancy has also been associated with lower educational attainment and lower paid employment (Currie et al., 2020).

The binge-drinking definition derived from ALSPAC questionnaires, namely ‘two pints of beer or more or equivalent on one occasion’ (roughly between 4-6 units depending on the alcohol concentration), includes a lower unit range that than current definition of binge drinking for women (six units on one occasion) (Department of Health and Social Care [hereafter DHSC], 2021). Given the findings within this study we may hypothesise that the observed associations between PAE and MRBs may be stronger at the current definition level as greater alcohol exposure would likely be more harmful to the developing foetus.

There are multiple biologically plausible and explanatory theories for associations between PAE and adolescent risk behaviour development. Alcohol is a known teratogen which can lead to abnormal brain development in utero (Chung et al. 2021). Maternal genotypes can also influence alcohol drinking behaviour and alcohol metabolism (Sambo and Goldman, 2023). These factors, coupled with certain infant genotypes thought to influence susceptibility to harmful PAE effects may influence the likelihood of FASD being developed (Gemma et al., 2007). It is also hypothesised that epigenetic processes such as alterations in DNA methylation and histone methylation, which influence gene expression, continue to influence brain development long after initial PAE (Jarmasz et al., 2019). Studies in rats, for example, have shown that prenatal alcohol exposure potentiates increased alcohol consumption in the offspring at adolescence (Fabio et al., 2015; Marengo et al., 2025).

### Study strengths and limitations

The key strengths of this study include the use of a large population-based cohort with prospective, detailed measurement of prenatal alcohol exposure. This enabled investigation of different patterns, doses (including low doses) and frequency of alcohol exposure on a range of MRB outcomes.

Given the range of MRB outcomes examined, using a q-value<0.05 threshold to account for false discovery rates, reduces the potential for Type 1 errors in our results. Associations detected at a p-value <0.05 but with attenuated q-values (≥0.05) - such as associations between infrequent PAE and adolescent hazardous alcohol use - may still reflect true effects. However, because multiple hypotheses were tested, the risk of Type I error is elevated and, compared to q-values, unadjusted p-values provide less certainty that these findings are not due to chance.

We must note that, as questionnaire responses for PAE were based on responses in trimesters 1 and 2 of pregnancy, we can draw no clear conclusions about the impacts of PAE before conception or during trimester 3. In addition, despite the use of a DAG to model variable relationships in the substantive model, we must be aware that residual confounding may exist that may influence associations between PAE and MRBs.

As there is also the potential for ongoing maternal/or paternal drinking and the clustering of these with other factors in the postnatal environment (e.g. other lifestyle factors), it is unclear whether the effects demonstrated in this study are directly due to the biological impacts of alcohol in-utero, or the effects of the postnatal environment. Future research may wish to use other causal inference techniques (e.g. paternal controls) to help elucidate the relative contribution of prenatal and postnatal factors on the outcomes we have observed in adolescents in this study.

Inclusion of polygenic scores for alcohol use should also be considered to help address residual confounding. A further option is to use a within-family study design comparing outcomes in siblings who were exposed to varying levels of PAE.

Attrition within the ALSPAC cohort has been shown to be associated with low socio-economic status (SES) leading to underestimation of the effects of inequalities (Howe et al., 2013). It is unclear what impacts this attrition would have had on our study, given that higher SES in our sample was more associated with regular, sustained prenatal alcohol exposure. However, given that low SES may be related to prenatal binge drinking exposure and hazardous alcohol use in adolescence, this may have led to underestimation of the impacts of prenatal binge drinking exposure on MRBs (Katikireddi et al., 2017). Given the neurobehavioral impacts of FASD, it could also be hypothesised that those most impacted by PAE may be those least likely to repeatedly respond to the questionnaires, leading to further attrition bias. This may be reflected in results through both a reduction in the strength and size of the association between PAE on multiple risk behaviour development. The use of multiple imputation in this study will have helped mitigate the impacts of selection bias due to missing data and will have improved precision in our estimates.

Underestimation of alcohol consumption is a known issue when using self-report measures. This has been demonstrated in studies that compare reported alcohol use in questionnaires with sales data (Stockwell et al., 2018). Considerable stigma experienced around the impacts of PAE may further impact information disclosure about both alcohol and wider substance use (Stone, 2015). The low thresholds set here for inclusion in the ‘infrequent’ PAE group (less than one glass of wine or equivalent per week) is both a strength and limitation, by capturing more of those underestimating their intake but also potentially leading to an overestimation of the impacts of lower reported levels of PAE.

When defining our exposure groups, we found that stratifying the ‘frequent’ PAE group into more categories was not deemed possible due to the rarity of higher levels of reported PAE leading to inadequate sample sizes for analysis. While potential effects of unit increases may have been explored, very few mothers reported drinking more than 2 units per week. As such, focusing on a stratified unit score may have resulted inadequately sized groups for analysis. There was also substantially more missingness in the continuous measure for PAE, than the selected PAE variables.

Finally, the use of a continuous outcome variable (the MRB score) to summarise occurrences of distinct risk behaviours, is useful in examining the overall number of risk behaviour differences between exposure groups. However, the overall score, particularly after model adjustment obfuscates the individual MRB components contributing to it. This continuous score also implicitly assumes that all MRBs can be considered equivalent. This represents an oversimplification in terms of health impacts. We addressed this by examining the likelihood of individual multiple risk behaviours occurring in those with different PAE profiles.

### Implications

This study supports current UK Chief Medical Officers (CMOs) guidance that the safest approach if pregnant, or planning a pregnancy, is to not drink alcohol.

Adolescents in our study had an increased risk of engaging in harmful alcohol use when exposed prenatally to the equivalent of one or more drinks of alcohol per week. In addition, adolescents exposed prenatally to binge-level alcohol exposure (here defined as two pints of beer or equivalent on one or more occasion) were at increased risk of engaging in underage sexual intercourse and had higher multiple risk behaviour scores than those who were unexposed to binge-level alcohol use prenatally. This is important as a higher number of developed multiple risk behaviours are associated with greater morbidity and mortality (Wright et al., 2018). These findings may have implications for how clinicians and public health professionals explain the potential long-term risks of PAE to parents who are pregnant or planning to become pregnant in the prevention or reduction of PAE and in how clinicians may screen for hazardous alcohol use in those with PAE later in life. It is crucial however that any opportunities healthcare professionals’ take to discuss alcohol intake, such as in pregnant mothers, are handled with utmost empathy and a non-judgemental manner (National Institute for Health and Care Excellence, 2015).

The findings of an association between PAE from binge drinking and engagement in risky sexual behaviour in adolescence (underage sexual intercourse), coupled with the findings of frequent PAE being associated with hazardous alcohol use at adolescence, warrant further research to explore the potential intergenerational impacts of PAE. To investigate this, research would need to look at how the prevalence of PAE changes in subsequent generations of those with PAE. Such research may have implications for targeted health education and prevention strategies for hazardous alcohol use coupled with sexually risky behaviours in adolescents with PAE.

## Data Availability

The informed consent obtained from ALSPAC participants does not allow the data to be made freely available through any third party maintained public repository. However, data used for this submission can be made available on request to the ALSPAC Executive. The ALSPAC data management plan describes in detail the policy regarding data sharing, which is through a system of managed open access. Full instructions for applying for data access can be found here: http://www.bristol.ac.uk/alspac/researchers/access/. The ALSPAC study website contains details of all the data that are available
(http://www.bristol.ac.uk/alspac/researchers/our-data/)

## ACKNOWLEDGMENTS

We are extremely grateful to all the families who took part in this study, the midwives for their help in recruiting them, and the whole ALSPAC team, which includes interviewers, computer and laboratory technicians, clerical workers, research scientists, volunteers, managers, receptionists and nurses.

