## Supplementary Material for "Prenatal alcohol exposure and the development of multiple risk behaviours in adolescence: A birth cohort study"

**Supplementary Table 1. Complete case sample characteristics table**

|  | **Frequency of prenatal alcohol exposure (PAE) for complete cases** | | | | | **Prenatal binge drinking exposure groups for complete cases** | | | |
| --- | --- | --- | --- | --- | --- | --- | --- | --- | --- |
| **Maternal, child or partner characteristics** | **No PAE (N=632)** | **Infrequent PAE (N=1096)** | **Frequent PAE (N=277)** | **Total (N=2005)** | **p-value^a^** | **No binge drinking (N=1774)** | **Binge drinking (N=231)** | **Total (N=2005)** | **p-value^a^** |
| **Maternal age at delivery (years)** |  |  |  |  | **< 0.001** |  |  |  | **0.987** |
| Mean (SD) | 28.6 (4.1) | 29.8 (4.2) | 31.2 (4.0) | 29.6 (4.2) |  | 29.6 (4.2) | 29.6 (4.3) | 29.6 (4.2) |  |
| Range | 17.0 - 41.0 | 17.0 - 42.0 | 21.0 - 44.0 | 17.0 - 44.0 |  | 17.0 - 44.0 | 20.0 - 41.0 | 17.0 - 44.0 |  |
| **Any smoking during pregnancy** |  |  |  |  | **< 0.001** |  |  |  | **< 0.001** |
| No | 571 (90.3%) | 960 (87.6%) | 226 (81.6%) | 1757 (87.6%) |  | 1584 (89.3%) | 173 (74.9%) | 1757 (87.6%) |  |
| Yes | 61 (9.7%) | 136 (12.4%) | 51 (18.4%) | 248 (12.4%) |  | 190 (10.7%) | 58 (25.1%) | 248 (12.4%) |  |
| **Maternal cannabis use during pregnancy** |  |  |  |  | **< 0.001** |  |  |  | **0.016** |
| No | * | 1082 (98.7%) | 262 (94.6%) | 1973 (98.4%) |  | 1750 (98.6%) | 223 (96.5%) | 1973 (98.4%) |  |
| Yes | * | 14 (1.3%) | 15 (5.4%) | 32 (1.6%) |  | 24 (1.4%) | 8 (3.5%) | 32 (1.6%) |  |
| **Maternal drug use (excluding cannabis) during pregnancy** |  |  |  |  | **0.486** |  |  |  | **0.398** |
| No | * | * | * | * |  | * | * | * |  |
| Yes | * | * | * | * |  | * | * | * |  |
| **Parity** |  |  |  |  | **< 0.001** |  |  |  | **0.204** |
| Nulliparous | 382 (60.4%) | 568 (51.8%) | 147 (53.1%) | 1097 (54.7%) |  | 976 (55.0%) | 121 (52.4%) | 1097 (54.7%) |  |
| Primiparous | 191 (30.2%) | 381 (34.8%) | 80 (28.9%) | 652 (32.5%) |  | 580 (32.7%) | 72 (31.2%) | 652 (32.5%) |  |
| Multiparous | 59 (9.3%) | 147 (13.4%) | 50 (18.1%) | 256 (12.8%) |  | 218 (12.3%) | 38 (16.5%) | 256 (12.8%) |  |
| **Maternal social class** |  |  |  |  | **0.031** |  |  |  | **0.034** |
| I to III (Non-manual) | 533 (84.3%) | 964 (88.0%) | 249 (89.9%) | 1746 (87.1%) |  | 1555 (87.7%) | 191 (82.7%) | 1746 (87.1%) |  |
| III (Manual) to V | 99 (15.7%) | 132 (12.0%) | 28 (10.1%) | 259 (12.9%) |  | 219 (12.3%) | 40 (17.3%) | 259 (12.9%) |  |
| **Mother’s highest educational qualification** |  |  |  |  | **< 0.001** |  |  |  | **0.012** |
| A-level or above | 297 (47.0%) | 634 (57.8%) | 178 (64.3%) | 1109 (55.3%) |  | 999 (56.3%) | 110 (47.6%) | 1109 (55.3%) |  |
| Lower than A-level | 335 (53.0%) | 462 (42.2%) | 99 (35.7%) | 896 (44.7%) |  | 775 (43.7%) | 121 (52.4%) | 896 (44.7%) |  |
| **Home ownership status** |  |  |  |  | **0.647** |  |  |  | **0.053** |
| Own home | 565 (89.4%) | 982 (89.6%) | 253 (91.3%) | 1800 (89.8%) |  | 1601 (90.2%) | 199 (86.1%) | 1800 (89.8%) |  |
| No owned home or mortgage | 67 (10.6%) | 114 (10.4%) | 24 (8.7%) | 205 (10.2%) |  | 173 (9.8%) | 32 (13.9%) | 205 (10.2%) |  |
| **Marital status** |  |  |  |  | **< 0.001** |  |  |  | **< 0.001** |
| Married | 563 (89.1%) | 955 (87.1%) | 221 (79.8%) | 1739 (86.7%) |  | 1562 (88.0%) | 177 (76.6%) | 1739 (86.7%) |  |
| Not married | 69 (10.9%) | 141 (12.9%) | 56 (20.2%) | 266 (13.3%) |  | 212 (12.0%) | 54 (23.4%) | 266 (13.3%) |  |
| **Sex of child** |  |  |  |  | **0.454** |  |  |  | **0.537** |
| Female | 387 (61.2%) | 647 (59.0%) | 158 (57.0%) | 1192 (59.5%) |  | 1059 (59.7%) | 133 (57.6%) | 1192 (59.5%) |  |
| Male | 245 (38.8%) | 449 (41.0%) | 119 (43.0%) | 813 (40.5%) |  | 715 (40.3%) | 98 (42.4%) | 813 (40.5%) |  |
| **Child birthweight** |  |  |  |  | **0.601** |  |  |  | **0.924** |
| Mean (SD) | 3405.4 (533.4) | 3429.1 (502.7) | 3434.6 (549.3) | 3422.4 (519.0) |  | 3422.0 (519.1) | 3425.4 (519.5) | 3422.3 (519.0) |  |
| Range | 910.0 - 5050.0 | 1040.0 - 4900.0 | 1090.0 - 5080.0 | 910.0 - 5080.0 |  | 910.0 - 5080.0 | 1049.0 - 4760.0 | 910.0 - 5080.0 |  |
| **Length of pregnancy (gestational length) (weeks)** |  |  |  |  | **0.387** |  |  |  | **0.962** |
| Extreme pre-term | 0 (0.0%) | 0 (0.0%) | 0 (0.0%) | 0 (0.0%) |  | 0 (0.0%) | 0 (0.0%) | 0 (0.0%) |  |
| Pre-term | 29 (4.6%) | 43 (3.9%) | 16 (5.8%) | 88 (4.4%) |  | 78 (4.4%) | 10 (4.3%) | 88 (4.4%) |  |
| Term | 603 (95.4%) | 1053 (96.1%) | 261 (94.2%) | 1917 (95.6%) |  | 1696 (95.6%) | 221 (95.7%) | 1917 (95.6%) |  |
| **Partner’s alcohol consumption during pregnancy** |  |  |  |  | **< 0.001** |  |  |  | **0.003** |
| Never | 33 (5.2%) | 18 (1.6%) | * | * |  | 51 (2.9%) | * | * |  |
| Infrequent | 179 (28.3%) | 215 (19.6%) | * | * |  | 377 (21.3%) | * | * |  |
| Frequent | 420 (66.5%) | 863 (78.7%) | 261 (94.2%) | 1544 (77.0%) |  | 1346 (75.9%) | 198 (85.7%) | 1544 (77.0%) |  |
| **MRB Score** |  |  |  |  | **0.004** |  |  |  | **0.002** |
| Mean (SD) | 1.11 (1.33) | 1.22 (1.35) | 1.43 (1.44) | 1.21 (1.36) |  | 1.18 (1.33) | 1.48 (1.54) | 1.21 (1.36) |  |
| Range | 0.0 - 6.0 | 0.0 - 6.0 | 0.0 - 6.0 | 0.0 - 6.0 |  | 0.0 - 6.0 | 0.0 - 6.0 | 0.0 - 6.0 |  |
| **MRB - Hazardous alcohol use** |  |  |  |  | **< 0.001** |  |  |  | **0.005** |
| No | 451 (71.4%) | 690 (63.0%) | 146 (52.7%) | 1287 (64.2%) |  | 1158 (65.3%) | 129 (55.8%) | 1287 (64.2%) |  |
| Yes | 181 (28.6%) | 406 (37.0%) | 131 (47.3%) | 718 (35.8%) |  | 616 (34.7%) | 102 (44.2%) | 718 (35.8%) |  |
| **MRB - Regular smoking** |  |  |  |  | **0.081** |  |  |  | **0.022** |
| No | 560 (88.6%) | 991 (90.4%) | 238 (85.9%) | 1789 (89.2%) |  | 1593 (89.8%) | 196 (84.8%) | 1789 (89.2%) |  |
| Yes | 72 (11.4%) | 105 (9.6%) | 39 (14.1%) | 216 (10.8%) |  | 181 (10.2%) | 35 (15.2%) | 216 (10.8%) |  |
| **MRB - Cannabis use** |  |  |  |  | **0.276** |  |  |  | **0.063** |
| No | 601 (95.1%) | 1030 (94.0%) | 256 (92.4%) | 1887 (94.1%) |  | 1605 (90.5%) | 200 (86.6%) | 1805 (90.0%) |  |
| Yes | 31 (4.9%) | 66 (6.0%) | 21 (7.6%) | 118 (5.9%) |  | 169 (9.5%) | 31 (13.4%) | 200 (10.0%) |  |
| **MRB - Drug use (excluding cannabis)** |  |  |  |  | **0.099** |  |  |  | **0.005** |
| No | 579 (91.6%) | 985 (89.9%) | 241 (87.0%) | 1805 (90.0%) |  | 1679 (94.6%) | 208 (90.0%) | 1887 (94.1%) |  |
| Yes | 53 (8.4%) | 111 (10.1%) | 36 (13.0%) | 200 (10.0%) |  | 95 (5.4%) | 23 (10.0%) | 118 (5.9%) |  |
| **MRB - Unsafe sex** |  |  |  |  | **0.657** |  |  |  | **0.998** |
| No | 623 (98.6%) | 1081 (98.6%) | * | * |  | 1751 (98.7%) | * | * |  |
| Yes | 9 (1.4%) | 15 (1.4%) | * | * |  | 23 (1.3%) | * | * |  |
| **MRB - Underage sex** |  |  |  |  | **0.797** |  |  |  | **0.055** |
| No | 541 (85.6%) | 949 (86.6%) | 241 (87.0%) | 1731 (86.3%) |  | 1541 (86.9%) | 190 (82.3%) | 1731 (86.3%) |  |
| Yes | 91 (14.4%) | 147 (13.4%) | 36 (13.0%) | 274 (13.7%) |  | 233 (13.1%) | 41 (17.7%) | 274 (13.7%) |  |
| **MRB - Antisocial behaviour** |  |  |  |  | **0.275** |  |  |  | **0.505** |
| No | 367 (58.1%) | 614 (56.0%) | 145 (52.3%) | 1126 (56.2%) |  | 1001 (56.4%) | 125 (54.1%) | 1126 (56.2%) |  |
| Yes | 265 (41.9%) | 482 (44.0%) | 132 (47.7%) | 879 (43.8%) |  | 773 (43.6%) | 106 (45.9%) | 879 (43.8%) |  |
| **Abbreviations: PAE = prenatal alcohol exposure, MRB = multiple risk behaviour** | | | | | |  |  |  |  |
| **^a^ P-values calculated using chi-squared analyses for categorical variables and ANOVA for continuous variables** | | | | | | | | |  |
| **The * sign indicates suppressed counts of <5 people, or counts where a supressed number can be calculated e.g. from the percentage of another count.** | | | | | | | | | |
| **Note: Percentages by covariate group may not sum to 100% due to rounding to 1 decimal place** | | | | | | |  |  |  |

**Supplementary Material Table 2. Characteristics of complete and incomplete cases**

| **Maternal, child or partner characteristics** | **Incomplete or missing cases (N=12598)** | **Complete cases (N=2005)** | **Total (N=14603)** | **p-value^a^** |
| --- | --- | --- | --- | --- |
| **Prenatal exposure to alcohol** |  |  |  | **< 0.001** |
| Missing | 1748 | 0 | 1748 |  |
| No | 4013 (37.0%) | 632 (31.5%) | 4645 (36.1%) |  |
| Infrequent | 5320 (49.0%) | 1096 (54.7%) | 6416 (49.9%) |  |
| Frequent | 1517 (14.0%) | 277 (13.8%) | 1794 (14.0%) |  |
| **Prenatal exposure to binge drinking** |  |  |  | **< 0.001** |
| Missing | 1733 | 0 | 1733 |  |
| No | 8908 (82.0%) | 1774 (88.5%) | 10682 (83.0%) |  |
| Yes | 1957 (18.0%) | 231 (11.5%) | 2188 (17.0%) |  |
| **Maternal age at delivery** |  |  |  | **< 0.001** |
| Missing | 716 | 0 | 716 |  |
| Mean (SD) | 27.7 (5.0) | 29.6 (4.2) | 28.0 (5.0) |  |
| Range | 15.0 - 44.0 | 17.0 - 44.0 | 15.0 - 44.0 |  |
| **Any smoking during pregnancy** |  |  |  | **< 0.001** |
| Missing | 1532 | 0 | 1532 |  |
| No | 8037 (72.6%) | 1757 (87.6%) | 9794 (74.9%) |  |
| Yes | 3029 (27.4%) | 248 (12.4%) | 3277 (25.1%) |  |
| **Maternal cannabis use during pregnancy** |  |  |  | **< 0.001** |
| Missing | 2234 | 0 | 2234 |  |
| No | 10048 (97.0%) | 1973 (98.4%) | 12021 (97.2%) |  |
| Yes | 316 (3.0%) | 32 (1.6%) | 348 (2.8%) |  |
| **Maternal drug use (excluding cannabis) during pregnancy** |  |  |  | **0.046** |
| Missing | 1777 | 0 | 1777 |  |
| No | 10763 (99.5%) | * | * |  |
| Yes | 58 (0.5%) | * | * |  |
| **Parity** |  |  |  | **< 0.001** |
| Missing | 1760 | 0 | 1760 |  |
| Nulliparous | 4636 (42.8%) | 1097 (54.7%) | 5733 (44.6%) |  |
| Primiparous | 3851 (35.5%) | 652 (32.5%) | 4503 (35.1%) |  |
| Multiparous | 2351 (21.7%) | 256 (12.8%) | 2607 (20.3%) |  |
| **Maternal social class** |  |  |  | **< 0.001** |
| Missing | 4609 | 0 | 4609 |  |
| I to III (Non-manual) | 6265 (78.4%) | 1746 (87.1%) | 8011 (80.2%) |  |
| III (Manual) to V | 1724 (21.6%) | 259 (12.9%) | 1983 (19.8%) |  |
| **Mother’s highest educational qualification** |  |  |  | **< 0.001** |
| Missing | 2273 | 0 | 2273 |  |
| A-level or above | 3247 (31.4%) | 1109 (55.3%) | 4356 (35.3%) |  |
| Lower than A-level | 7078 (68.6%) | 896 (44.7%) | 7974 (64.7%) |  |
| **Home ownership status** |  |  |  | **< 0.001** |
| Missing | 1660 | 0 | 1660 |  |
| Own home | 7696 (70.4%) | 1800 (89.8%) | 9496 (73.4%) |  |
| No owned home or mortgage | 3242 (29.6%) | 205 (10.2%) | 3447 (26.6%) |  |
| **Marital status** |  |  |  | **< 0.001** |
| Missing | 1601 | 0 | 1601 |  |
| Married | 8004 (72.8%) | 1739 (86.7%) | 9743 (74.9%) |  |
| Not married | 2993 (27.2%) | 266 (13.3%) | 3259 (25.1%) |  |
| **Sex of child** |  |  |  | **< 0.001** |
| Missing | 0 | 0 | 0 |  |
| Female | 5932 (47.1%) | 1192 (59.5%) | 7124 (48.8%) |  |
| Male | 6666 (52.9%) | 813 (40.5%) | 7479 (51.2%) |  |
| **Child birthweight** |  |  |  | **0.008** |
| Missing | 895 | 0 | 895 |  |
| Mean (SD) | 3386.5 (565.2) | 3422.4 (519.0) | 3391.8 (558.8) |  |
| Range | 645.0 – 5640.0 | 910.0 0 5080.0 | 645.0 – 5640.0 |  |
| **Length of pregnancy (gestational length) (weeks)** |  |  |  | **0.002** |
| Missing | 716 | 0 | 716 |  |
| Extreme pre-term | 10 (0.1%) | 0 (0.0%) | 10 (0.1%) |  |
| Pre-term | 739 (6.2%) | 88 (4.4%) | 827 (6.0%) |  |
| Term | 11133 (93.7%) | 1917 (95.6%) | 13050 (94.0%) |  |
| **Partner’s alcohol consumption during pregnancy** |  |  |  | **< 0.001** |
| Missing | 4964 | 0 | 4964 |  |
| Never | 423 (5.5%) | 54 (2.7%) | 477 (4.9%) |  |
| Infrequent | 1955 (25.6%) | 407 (20.3%) | 2362 (24.5%) |  |
| Frequent | 5256 (68.8%) | 1544 (77.0%) | 6800 (70.5%) |  |
| **Hazardous alcohol consumption at age 16** |  |  |  | **0.093** |
| Missing | 10077 | 0 | 10077 |  |
| No | 1557 (61.8%) | 1287 (64.2%) | 2844 (62.8%) |  |
| Yes | 964 (38.2%) | 718 (35.8%) | 1682 (37.2%) |  |
| **Regular smoking at age 16** |  |  |  | **0.005** |
| Missing | 9567 | 0 | 9567 |  |
| No | 2623 (86.5%) | 1789 (89.2%) | 4412 (87.6%) |  |
| Yes | 408 (13.5%) | 216 (10.8%) | 624 (12.4%) |  |
| **Regular cannabis use at age 16** |  |  |  | **0.828** |
| Missing | 9564 | 0 | 9564 |  |
| No | 2737 (90.2%) | 1805 (90.0%) | 4542 (90.1%) |  |
| Yes | 297 (9.8%) | 200 (10.0%) | 497 (9.9%) |  |
| **Drug use at age 16** |  |  |  | **0.939** |
| Missing | 9769 | 0 | 9769 |  |
| No | 2664 (94.2%) | 1887 (94.1%) | 4551 (94.1%) |  |
| Yes | 165 (5.8%) | 118 (5.9%) | 283 (5.9%) |  |
| **Criminal or antisocial behaviour by age 16** |  |  |  | **0.005** |
| Missing | 9264 | 0 | 9264 |  |
| No | 1740 (52.2%) | 1126 (56.2%) | 2866 (53.7%) |  |
| Yes | 1594 (47.8%) | 879 (43.8%) | 2473 (46.3%) |  |
| **Sexual intercourse without contraception in last year by age 16** |  |  |  | **0.208** |
| Missing | 9444 | 0 | 9444 |  |
| No | 3099 (98.3%) | 1979 (98.7%) | 5078 (98.4%) |  |
| Yes | 55 (1.7%) | 26 (1.3%) | 81 (1.6%) |  |
| **Sexual intercourse before 16th birthday** |  |  |  | **< 0.001** |
| Missing | 9441 | 0 | 9441 |  |
| No | 2569 (81.4%) | 1731 (86.3%) | 4300 (83.3%) |  |
| Yes | 588 (18.6%) | 274 (13.7%) | 862 (16.7%) |  |
| **^a^ P-values calculated using chi-squared analyses for categorical variables and ANOVA for continuous variables**  **The * sign indicates suppressed counts of <5 people, or counts where a suppressed number can be calculated e.g. from the total of another count.**  **Note: Percentages by covariate group may not sum to 100% due to rounding to 1 decimal place** | | | | |

**Supplementary Table 3. Complete case strategy results (n=2,005) comparing overall MRB score between PAE exposure and comparator groups**

| **PAE exposure group** | **Unadjusted or adjusted** | **Coefficient/MRB Score difference (95% CI)** | **p-value** | **q-value (if p<0.05)** |
| --- | --- | --- | --- | --- |
| Infrequent PAE  (compared with no alcohol) | Unadjusted | (+)0.10 (-0.03 to +0.24) | 0.12 | - |
|  | Adjusted | (+)0.06 (-0.08 to +0.19) | 0.41 | - |
| Frequent PAE  (compared with no alcohol) | Unadjusted | (+)0.32 (+0.13 to +0.51) | <0.001 | 0.01 |
|  | Adjusted | (+)0.19 (-0.01 to +0.39) | 0.07 | - |
| Binge drinking  (compared with no binge drinking) | Unadjusted | (+)0.30 (+0.11 to +0.48) | 0.002 | 0.02 |
|  | Adjusted | (+)0.16 (-0.03 to +0.35) | 0.09 | - |

**Supplementary Table 4. Complete case strategy results (n=2,005) showing the odds ratios of individual MRBs occurring in PAE exposure groups relative to comparator groups**

|  |  | **Pre-natal alcohol exposure** | | | | | | | | |
| --- | --- | --- | --- | --- | --- | --- | --- | --- | --- | --- |
| **Multiple risk behaviour at age sixteen** | **Unadjusted or adjusted** | **Infrequent alcohol (compared with no alcohol)** | | | **Frequent alcohol (compared with no alcohol)** | | | **Binge drinking (compared with no binge drinking)** | | |
|  |  | **Odds ratio (95% CI)** | **p-value** | **q-value (if p<0.05)** | **Odds ratio (95% CI)** | **p-value** | **q-value (if p<0.05)** | **Odds ratio (95% CI)** | **p-value** | **q-value (if p<0.05)** |
| Hazardous alcohol use | Unadjusted | 1.47 (1.19-1.81) | <0.001 | 0.006 | 2.24 (1.67-3.00) | <0.001 | <0.001 | 1.49 (1.12-1.96) | 0.005 | 0.04 |
|  | Adjusted | 1.35 (1.09-1.68) | 0.007 | 0.04 | 1.89 (1.38-2.58) | <0.001 | 0.001 | 1.26 (0.94-1.68) | 0.11 | - |
| Regular smoking | Unadjusted | 0.82 (0.60-1.14) | 0.23 | - | 1.27 (0.83-1.93) | 0.26 | - | 1.57 (1.05-2.30) | 0.02 | 0.11 |
|  | Adjusted | 0.82 (0.59-1.15) | 0.25 | - | 1.24 (0.78-1.94) | 0.35 | - | 1.30 (0.85-1.93) | 0.21 | - |
| Regular cannabis use | Unadjusted | 1.23 (0.88-1.75) | 0.23 | - | 1.63 (1.03-2.55) | 0.03 | 0.14 | 1.47 (0.96-2.19) | 0.06 | - |
|  | Adjusted | 1.02 (0.72-1.47) | 0.89 | - | 1.05 (0.65-1.70) | 0.83 | - | 1.22 (0.78-1.85) | 0.37 | - |
| Drug use | Unadjusted | 1.24 (0.81-1.95) | 0.33 | - | 1.59 (0.89-2.80) | 0.11 | - | 1.95 (1.19-3.10) | 0.006 | 0.04 |
|  | Adjusted | 1.12 (0.72-1.77) | 0.63 | - | 1.20 (0.64-2.20) | 0.56 | - | 1.78 (1.06-2.87) | 0.02 | 0.11 |
| Antisocial behaviour | Unadjusted | 1.09 (0.89-1.33) | 0.41 | - | 1.26 (0.95-1.67) | 0.11 | - | 1.10 (0.83-1.45) | 0.51 | - |
|  | Adjusted | 1.05 (0.86-1.29) | 0.63 | - | 1.11 (0.82-1.51) | 0.48 | - | 0.99 (0.74-1.31) | 0.93 | - |
| Unsafe sex | Unadjusted | 0.96 (0.42-2.30) | 0.92 | - | 0.50 (0.07-1.97) | 0.38 | - | 1.00 (0.24-2.91) | 0.998 | - |
|  | Adjusted | 0.97 (0.42-2.38) | 0.94 | - | 0.51 (0.07-2.15) | 0.41 | - | 0.78 (0.18-2.36) | 0.70 | - |
| Underage sex | Unadjusted | 0.92 (0.70-1.22) | 0.57 | - | 0.89 (0.58-1.33) | 0.58 | - | 1.42 (0.98-2.04) | 0.06 | - |
|  | Adjusted | 0.93 (0.69-1.25) | 0.63 | - | 0.86 (0.55-1.34) | 0.51 | - | 1.19 (0.80-1.72) | 0.38 | - |
| Adjustment model covariates were maternal age, parity, birthweight, maternal social class, marital status, home ownership status, highest maternal educational qualification, maternal smoking, maternal cannabis use, maternal drug use (excluding cannabis), and paternal alcohol use during pregnancy. | | | | | | | | | | |

**Multiple imputation – additional information**

Through systematically appraising the ALSPAC variable catalogue, a wide range of variables were selected which were considered to help explain missingness in substantive model variables. The ‘quickpred-pt2’ method was then used to assign variables to the imputation model with a correlation cut off (‘mincor’) of 0.2. Two samples were imputed, a full sample for all records in the ALSPAC data (N = 14,603) and a partial imputation step was performed with substantially less imputed data, where individuals with a response to at least 1 MRB variable have been included (N = 6,752). For each sample, two multiple imputation analyses were performed. One for calculation of the MRB score outcome(where the total number of MRBs per individual is the outcome in the substantive model) and one for the individual MRB component analysis (where individual MRBs are the outcomes in the substantive model). The MRB score outcome imputation was passively imputed by calculating the sum of the imputed individual MRB components. For the full imputation, 70 imputations were performed (corresponding to roughly 70% of missing outcome data) across 150 iterations for the MRB score analysis and 50 iterations for the individual MRB logistic regression analysis for stability. For partial imputation (the primary presented analysis), given the smaller degree of missingness, 80 iterations and 70 imputations were performed to achieve stability. The component-based logistic regression analysis of MRBs was carried out with 50 iterations for 50 imputations steps.

All estimates for the MRB score (linear regression) and individual MRB components (odds ratios) were derived from pooled multiple imputation estimates using Rubin’s rules. Overall, the results did not substantially differ between our full and partial imputations strategies. Supplementary tables 5 and 6 show the sample characteristics for the full imputation, where n=14,603 records were imputed. Supplementary tables 7 and 8 show the results from the full imputation. Supplementary table 9 shows degree of missingness in the full imputation model. Diagnostics were performed on the full and partial imputation step with mean and standard deviation graphs across iterations to show the stability of the imputations (Supplementary Figures A-P).

**Supplementary table 5. Multiple imputation sample characteristics table for the component-based MRB analysis for full imputation (N = 14,603)**

|  | **Frequency of prenatal alcohol exposure (PAE)** | | | | | **Prenatal binge drinking exposure groups** | | | |
| --- | --- | --- | --- | --- | --- | --- | --- | --- | --- |
| **Maternal, child or partner characteristics** | **No PAE (N=5,332)** | **Infrequent PAE (N=7,247)** | **Frequent PAE (N=2025)** | **Total (N=14,603)** | **p-value^a^** | **No binge drinking (N=12,111)** | **Binge drinking (N=2,492)** | **Total (N=14,603)** | **p-value^a^** |
| **Maternal age at delivery (years)** |  |  |  |  | **<0.001** |  |  |  | **0.647** |
| ≥19 years | 337 (6.3%) | 269 (3.7%) | 71 (3.5%) | 677 (4.6%) |  | 560 (4.6%) | 116 (4.7%) | 677 (4.6%) |  |
| >19 years and ≤ 34 years | 4598 (86.2%) | 6234 (86.0%) | 1644 (81.2%) | 12,476 (85.4%) |  | 10,360 (85.5%) | 2,116 (84.9%) | 12,476 (85.4%) |  |
| ≥35 years | 397 (7.4%) | 744 (10.3%) | 309 (15.3%) | 1450 (9.9%) |  | 1,190 (9.8%) | 260 (10.4%) | 1,450 (9.9%) |  |
| **Any smoking during pregnancy** |  |  |  |  | **<0.001** |  |  |  | **<0.001** |
| No | 4058 (76.1%) | 5482 (75.6%) | 1,344 (66.4%) | 10,884 (74.5%) |  | 9,408 (77.7%) | 1,476 (59.2%) | 10,884 (74.5%) |  |
| Yes | 1274 (23.9%) | 1,765 (24.4%) | 680 (33.6%) | 3,719 (25.5%) |  | 2,702 (22.3%) | 1,017 (40.8%) | 3,719 (25.5%) |  |
| **Maternal cannabis use during pregnancy** |  |  |  |  | **<0.001** |  |  |  | **<0.001** |
| No | 5,255 (98.6%) | 7,032 (97.0%) | 1,906 (94.2%) | 14,193 (97.2%) |  | 11,849 (97.8%) | 2,343 (94.0%) | 14,193 (97.2%) |  |
| Yes | 77 (1.4%) | 215 (3.0%) | 118 (5.8%) | 410 (2.8%) |  | 261 (2.2%) | 149 (6.0%) | 410 (2.8%) |  |
| **Maternal drug use (excluding cannabis) during pregnancy** |  |  |  |  | **<0.001** |  |  |  | **<0.001** |
| No | 5,315 (99.7%) | 7,210 (99.5%) | 2003 (99.0%) | 14,528 (99.5%) |  | 12,070 (99.7%) | 2,458 (98.6%) | 14,528 (99.5%) |  |
| Yes | 17 (0.3%) | 37 (0.5%) | 21 (1.0%) | 75 (0.5%) |  | 41 (0.3%) | 34 (1.4%) | 75 (0.5%) |  |
| **Parity** |  |  |  |  | **<0.001** |  |  |  | **<0.001** |
| Nulliparous | 2545 (47.7%) | 3,128 (43.2%) | 851 (42.0%) | 6,524 (44.7%) |  | 5,531 (45.7%) | 993 (39.8%) | 6,524 (44.6%) |  |
| Primiparous | 1782 (33.4%) | 2,633 (36.3%) | 677 (33.5%) | 5092 (34.9%) |  | 4247 (35.1%) | 845 (33.9%) | 5092 (34.9%) |  |
| Multiparous | 1,005 (18.8%) | 1,486 (20.5%) | 496 (24.5%) | 2,987 (20.5%) |  | 2,333 (19.3%) | 655 (26.3%) | 2,987 (20.5%) |  |
| **Maternal social class** |  |  |  |  | **<0.001** |  |  |  | **<0.001** |
| I to III (Non-manual) | 3,906 (73.3%) | 5,687 (78.5%) | 1603 (79.2%) | 11,196 (76.7%) |  | 9,439 (77.9%) | 1,757 (70.5%) | 11,196 (76.7%) |  |
| III (Manual) to V | 1,425 (26.7%) | 1,560 (21.5%) | 422 (20.8%) | 3,407 (23.3%) |  | 2,672 (22.1%) | 736 (29.5%) | 3,407 (23.3%) |  |
| **Mother’s highest educational qualification** |  |  |  |  | **<0.001** |  |  |  | **<0.001** |
| A-level or above | 1,500 (28.1%) | 2,698 (37.2%) | 804 (39.7%) | 5,002 (34.3%) |  | 4,331 (35.8%) | 671 (26.9%) | 5,002 (34.3%) |  |
| Lower than A-level | 3832 (71.9%) | 4548 (62.8%) | 1,221 (60.3%) | 9,601 (65.7%) |  | 7,780 (64.2%) | 1,822 (73.1%) | 9,601 (65.7%) |  |
| **Home ownership status** |  |  |  |  | **<0.001** |  |  |  | **<0.001** |
| Own home | 3692 (69.3%) | 5,486 (75.7%) | 1,464 (72.3%) | 10,643 (72.9%) |  | 9,033 (74.6%) | 1,610 (64.6%) | 10,643 (72.9%) |  |
| No owned home or mortgage | 1,639 (30.7%) | 1,761 (24.3%) | 560 (27.7%) | 3,960 (27.1%) |  | 3,078 (25.4%) | 883 (35.4%) | 3,960 (27.1%) |  |
| **Marital status** |  |  |  |  | **<0.001** |  |  |  | **<0.001** |
| Married | 3,938 (73.9%) | 5,534 (76.4%) | 1,385 (68.4%) | 10,857 (74.3%) |  | 9,263 (76.5%) | 1,594 (63.9%) | 10,857 (74.3%) |  |
| Not married | 1,394 (26.1%) | 1,713 (23.6%) | 639 (31.6%) | 3,746 (25.7%) |  | 2,847 (23.5%) | 899 (36.1%) | 3,746 (25.7%) |  |
| **Sex of child** |  |  |  |  | **0.496** |  |  |  | **0.991** |
| Female | 2,610 (49.0%) | 3,551 (49.0%) | 963 (47.6%) | 7,124 (48.8%) |  | 5,909 (48.8%) | 1,215 (48.8%) | 7,124 (48.8%) |  |
| Male | 2,722 (51%) | 3,696 (51.0%) | 1,062 (52.4%) | 7479 (51.2%) |  | 6,202 (51.2%) | 1,277 (51.2%) | 7,479 (51.2%) |  |
| **Child birthweight** |  |  |  |  | **<0.001** |  |  |  | **0.842** |
| ≥2500 grams | 4,994 (93.7%) | 6,905 (95.3%) | 1,921 (94.9%) | 13,820 (94.6%) |  | 11458 (94.6%) | 2361 (94.7%) | 13,820 (94.6%) |  |
| <2,500 grams | 338 (6.3%) | 342 (4.7%) | 103 (5.1%) | 783 (5.4%) |  | 652 (5.4%) | 131 (5.3%) | 783 (5.4%) |  |
| **Length of pregnancy (gestational length)** |  |  |  |  | **0.005** |  |  |  | **0.371** |
| Extreme pre-term | * | * | * | 11 (0.1 %) |  | * | * | 11 (0.1%) |  |
| Pre-term | * | * | * | 870 (6.0%) |  | * | * | 870 (6.0%) |  |
| Term | 4,959 (93.0%) | 6,854 (94.6%) | 1,909 (94.3%) | 13,722 (94.0%) |  | 11,376 (93.9%) | 2,346 (94.1%) | 13,722 (94.0%) |  |
| **Partner’s alcohol consumption during pregnancy** |  |  |  |  | **<0.001** |  |  |  | **<0.001** |
| Never | 574 (10.8%) | 170 (2.3%) | 29 (1.4%) | 773 (5.3%) |  | 735 (6.1%) | 38 (1.5%) | 773 (5.3%) |  |
| Infrequent | 1,708 (32.0%) | 1,769 (24.4%) | 189 (9.3%) | 3,666 (25.1%) |  | 3,209 (26.5%) | 458 (18.4%) | 3666 (25.1%) |  |
| Frequent | 3,049 (57.2%) | 5,308 (73.2%) | 1,807 (89.2%) | 10,164 (69.6%) |  | 8,167 (67.4%) | 1,997 (80.1%) | 10,164 (69.6%) |  |
| **MRB - Hazardous alcohol use** |  |  |  |  | **<0.001** |  |  |  | **<0.001** |
| No | 3,491 (65.5%) | 4,320 (59.6%) | 1,039 (51.3%) | 8849 (60.6%) |  | 7,523 (62.1%) | 1,326 (53.2%) | 8849 (60.6%) |  |
| Yes | 1,841 (34.5%) | 2927 (40.4%) | 986 (48.7%) | 5754 (39.4%) |  | 4,588 (37.9%) | 1,166 (46.8%) | 5754 (39.4%) |  |
| **MRB - Drug use (excluding cannabis)** |  |  |  |  | **<0.001** |  |  |  | **<0.001** |
| No | 4,958 (93.0%) | 6,708 (92.6%) | 1,828 (90.3%) | 13,494 (92.4%) |  | 11,269 (93.1%) | 2,224 (89.2%) | 13,494 (92.4%) |  |
| Yes | 373 (7.0%) | 539 (7.4%) | 197 (9.7%) | 1,109 (7.6%) |  | 841 (6.9%) | 268 (10.8%) | 1,109 (7.6%) |  |
| **MRB - Cannabis use** |  |  |  |  | **<0.001** |  |  |  | **<0.001** |
| No | 4,806 (90.1%) | 6,345 (87.6%) | 1,727 (85.3%) | 12,878 (88.2%) |  | 10,764 (88.9%) | 2,114 (84.8%) | 12,878 (88.2%) |  |
| Yes | 525 (9.9%) | 902 (12.4%) | 298 (14.7%) | 1,725 (11.8%) |  | 1,346 (11.1%) | 379 (15.2%) | 1,725 (11.8%) |  |
| **MRB - Regular smoking** |  |  |  |  | **<0.001** |  |  |  | **<0.001** |
| No | 4,449 (83.4%) | 6,077 (83.9%) | 1,625 (80.3%) | 12,151 (83.2%) |  | 10,247 (84.6%) | 1,904 (76.4%) | 12,151 (83.2%) |  |
| Yes | 882 (16.6%) | 1,170 (16.1%) | 399 (19.7%) | 2,452 (16.8%) |  | 1,864 (15.4%) | 588 (23.6%) | 2,452 (16.8%) |  |
| **MRB - Antisocial behaviour** |  |  |  |  | **0.021** |  |  |  | **<0.001** |
| No | 2,805 (52.6%) | 3,664 (50.6%) | 1,002 (49.5%) | 7,471 (51.2%) |  | 6,344 (52.4%) | 1,127 (45.2%) | 7,471 (51.2%) |  |
| Yes | 2,527 (47.4%) | 3,583 (49.4%) | 1,023 (50.5%) | 7,132 (48.8%) |  | 5,767 (47.6%) | 1,365 (54.8%) | 7,132 (48.8%) |  |
| **MRB - Unsafe sex** |  |  |  |  | **0.411** |  |  |  | **<0.001** |
| No | 5,185 (97.3%) | 7,062 (97.5%) | 1,980 (97.8%) | 14,227 (97.4%) |  | 11,826 (97.7%) | 2,401 (96.3%) | 14,227 (97.4%) |  |
| Yes | 146 (2.7%) | 185 (2.5%) | 44 (2.2%) | 375 (2.6%) |  | 284 (2.3%) | 91 (3.7%) | 375 (2.6%) |  |
| **MRB - Underage sex** |  |  |  |  | **0.423** |  |  |  | **<0.001** |
| No | 4298 (80.6%) | 5,822 (80.3%) | 1,605 (79.3%) | 11,725 (80.3%) |  | 9,900 (81.7%) | 1,825 (73.2%) | 11,725 (80.3%) |  |
| Yes | 1,034 (19.4%) | 1,424 (19.7%) | 420 (20.7%) | 2,878 (19.7%) |  | 2,211 (18.3%) | 667 (26.8%) | 2,878 (19.7%) |  |
| **Abbreviations: PAE = prenatal alcohol exposure, MRB = multiple risk behaviour** | | | | | | | | | |
| **^a^ P-values calculated using chi-squared analyses for categorical variables.** | | | | | | | | | |
| **The * sign indicates suppressed counts of <5 people, or counts where a supressed number can be calculated e.g. from the percentage of another count.** | | | | | | | | | |
| **Participant counts may not sum to the row total N nor the column header total N, as values are based on pooled multiple imputation estimates rounded to the nearest integer. Column header totals are also rounded imputed values with the exception of the sample total N=14,603.** | | | | | | | | | |
| **Percentages by covariate or MRB group may not total 100% due to percentage rounding to 1 decimal place.** | | | | | | | | | |

**Supplementary table 6.** **Descriptive statistics of passively imputed adolescent MRB scores by prenatal alcohol exposure groups for full imputation (n=14,603)**

|  | **Frequency of prenatal alcohol exposure (PAE)** | | | | | **Prenatal binge drinking exposure groups** | | | |
| --- | --- | --- | --- | --- | --- | --- | --- | --- | --- |
| **Child characteristics** | **No PAE (N=5,334)** | **Infrequent PAE (N=7,249)** | **Frequent PAE (N=2,021)** | **Total (N=14,603)** | **p-value^a^** | **No binge drinking (N=12,111)** | **Binge drinking (N=2,492)** | **Total (N=14,603)** | **p-value^a^** |
| **MRB Score** |  |  |  |  | **<0.001** |  |  |  | **<0.001** |
| 0 | 2,856 (53.6%) | 3,566 (49.2%) | 902 (44.6%) | 7324 (50.2%) |  | 6,185 (51.1%) | 1,139 (45.7%) | 7324 (50.2%) |  |
| 1 | 710 (13.3%) | 1,112 (15.3%) | 296 (14.6%) | 2,118 (14.5%) |  | 1,797 (14.8%) | 321 (12.9%) | 2,118 (14.5%) |  |
| 2 | 581 (10.9%) | 951 (13.1%) | 304 (15%) | 1,835 (12.6%) |  | 1,521 (12.6%) | 314 (12.6%) | 1,835 (12.6%) |  |
| 3 | 301 (5.6%) | 507 (7.0%) | 167 (8.3%) | 975 (6.7%) |  | 798 (6.6%) | 177 (7.1%) | 975 (6.7%) |  |
| 4 | 196 (3.7%) | 329 (4.5%) | 104 (5.2%) | 629 (4.3%) |  | 500 (4.1%) | 129 (5.2%) | 629 (4.3%) |  |
| 5 | 210 (3.9%) | 291 (4.0%) | 98 (4.9%) | 599 (4.1%) |  | 457 (3.8%) | 142 (5.7%) | 599 (4.1%) |  |
| 6 | 283 (5.3%) | 317 (4.4%) | 117 (5.8%) | 717 (4.9%) |  | 535 (4.4%) | 182 (7.3%) | 717 (4.9%) |  |
| 7 | 197 (3.7%) | 176 (2.4%) | 33 (1.6%) | 406 (2.8%) |  | 319 (2.6%) | 87 (3.5%) | 406 (2.8%) |  |
| **Abbreviations: PAE = prenatal alcohol exposure, MRB = multiple risk behaviour.** | | | | | | | | | |
| **^a^ P-values calculated using chi-squared analysis for categorical (ordinal) variables.** | | | | | | | | | |
| **Participant totals may slightly differ from Table 3, as they are estimates derived from separate imputation models leading to inherent variability.** | | | | | | | | | |
| **Participant counts of MRB scores may not sum to the row total N nor the column header total N, as values are based on pooled multiple imputation estimates rounded to the nearest integer. Column header totals also rounded imputed values with the exception of the sample total N=14,603.** | | | | | | | | | |
| **Percentages by covariate or MRB group may not total 100% due to percentage rounding to 1 decimal place.** | | | | | | | | | |

**Supplementary Table 7. Full multiple imputation results (n=14,603) comparing adjusted MRB score between PAE exposure and comparator groups**

| **PAE exposure group** | **Adjusted MRB score difference (coefficient) (95% CI)** | **p-value** | **q-value (if p<0.05)** |
| --- | --- | --- | --- |
| Infrequent PAE  (compared with no alcohol) | (+)0.03 (-0.07 to +0.15) | 0.51 | - |
| Frequent PAE  (compared with no alcohol) | (+)0.13 (-0.04 to +0.30) | 0.13 | - |
| Binge drinking  (compared with no binge drinking) | (+)0.20 (+0.05 to +0.36) | 0.01 | 0.09 |

**Supplementary Table 8. Full multiple impution (n=14,603) aORs of MRBs in PAE exposure groups relative to comparator groups**

| **Multiple risk behaviour at age sixteen** | **Infrequent PAE (compared with no PAE)** | | | **Frequent PAE (compared with no PAE)** | | | **Binge drinking (compared to no binge drinking)** | | |
| --- | --- | --- | --- | --- | --- | --- | --- | --- | --- |
|  | **Adjusted odds ratio (95% CI)** | **p-value** | **q-value (if p<0.05)** | **Adjusted odds ratio (95% CI)** | **p-value** | **q-value (if p<0.05)** | **Adjusted odds ratio (95% CI)** | **p-value** | **q-value (if p<0.05)** |
| Hazardous alcohol use | 1.18 (1.02-1.35) | 0.02 | 0.14 | 1.43 (1.18-1.74) | <0.001 | 0.01 | 1.17 (1.01-1.35) | 0.04 | 0.19 |
| Regular smoking | 0.98 (0.83-1.17) | 0.88 | - | 1.12 (0.87-1.45) | 0.37 | - | 1.27 (0.99-1.64) | 0.06 | - |
| Regular cannabis use | 1.24 (0.98-1.57) | 0.07 | - | 1.26 (0.92-1.73) | 0.15 | - | 1.13 (0.84-1.51) | 0.43 | - |
| Drug use | 1.05 (0.80-1.40) | 0.69 | - | 1.20 (0.82-1.76) | 0.35 | - | 1.28 (0.90-1.81) | 0.17 | - |
| Antisocial behaviour | 1.09 (0.96-1.26) | 0.17 | - | 1.06 (0.88-1.27) | 0.54 | - | 1.16 (0.98-1.38) | 0.09 | - |
| Unsafe sexual intercourse | 0.97 (0.62-1.52) | 0.88 | - | 0.78 (0.38-1.62) | 0.50 | - | 1.29 (0.72-2.32) | 0.38 | - |
| Underage sexual intercourse | 1.00 (0.84-1.20) | 0.94 | - | 0.99 (0.79-1.25) | 0.94 | - | 1.35 (1.09-1.67) | 0.006 | 0.07 |
| Adjustment model covariates were maternal age, parity, birthweight, maternal social class, marital status, home ownership status, highest maternal educational qualification, maternal smoking, maternal cannabis use, maternal drug use (excluding cannabis), and paternal alcohol use during pregnancy. | | | | | | | | | |

**Supplementary Table 9. Imputed variables, variable allocation and imputation methods**

| **Variable type** | **Variable name** | **Maternal, paternal or child variable** | **Number of missing records imputed (percentage of sample imputed within sample with at least 1 MRB reported n=6,752)** | **Number of missing records imputed (percentage of sample imputed within total sample n=14,603)** | **Auxiliary variables assigned in MICE model using Quickpred-pt2 for partial imputation (in addition to the substantive model)** | **Auxiliary variables assigned in MICE model using Quickpred-pt2 for full imputation (in addition to the substantive model)** | **Imputation method for analysis 1 (MRB score outcome)** | **Imputation method for analysis 2 (Individual MRBs)** |
| --- | --- | --- | --- | --- | --- | --- | --- | --- |
| Confounder | Birthweight | Child | 392 (5.8%) | 895 (6.1%) | Correlated variables related to missingness are in the substantive model | Maternal physical activity, financial difficulties during pregnancy, crowding index, paternal social class | logreg | logreg |
| Confounder | Maternal age | Maternal | 311 (4.6%) | 716 (4.9%) | Correlated variables related to missingness are in the substantive model | *Correlated variables related to missingness are in the substantive model | polyreg | polyreg |
| Confounder | Gestation age | Maternal | 311 (4.6%) | 716 (4.9%) | Correlated variables related to missingness are in the substantive model | *Correlated variables related to missingness are in the substantive model | polyreg | polyreg |
| Confounder | Maternal smoking | Maternal | 437 (6.5%) | 1,532 (10.5%) | Correlated variables related to missingness are in the substantive model | Crowding index, family adversity during pregnancy | logreg | logreg |
| Confounder | Maternal cannabis use | Maternal | 692 (10.2%) | 2,234 (15.3%) | ACE parental substance use | Maternal self-harm, ACE parental substance use | logreg | logreg |
| Confounder | Maternal drug use | Maternal | 496 (7.3%) | 1,777 (12.2%) | Correlated variables related to missingness are in the substantive model | Alcohol binges in the last month, family adversity during pregnancy | logreg | logreg |
| Confounder | Parity | Maternal | 519 (7.7%) | 1,760 (12.1%) | Crowding index | Crown crisp depression score, physical activity, crowding index | polyreg | polyreg |
| Confounder | Maternal social class | Maternal | 1,341 (19.9%) | 4,609 (31.6%) | Maternal change in alcohol use, paternal social class, crowding index | Maternal change in alcohol use during pregnancy, paternal social class, crowding index | logreg | logreg |
| Confounder | Mother's highest educational qualification | Maternal | 529 (7.8%) | 2,273 (15.6%) | Paternal social class | Paternal social class, crowding index, ACE physical abuse, ACE emotional abuse, ACE parental offence, ACE parental mental health, ACE parental separation, child self-harm | logreg | logreg |
| Confounder | Home ownership status | Maternal | 524 (7.8%) | 1,660 (11.4%) | Paternal social class,  Crowding, Family adversity in pregnancy, family adversity at 0 to 2 years post-partum | Financial difficulties during pregnancy, maternal self-harm, crowding index, neighbourhood index, SDQ depression, SDQ hyperactivity, SDQ emotional symptoms, SDQ Peer problems, SDQ conduct problems, SDQ child total problems, ACE physical abuse, ACE sexual abuse, ACE emotional abuse, ACE bullying, ACE parental violence, ACE parental substance use, ACE parental mental health, ACE parental offence, ACE parental separation, ACE Neighbourhood, antisocial activities, sensation seeking score A, sensation seeking score B, family adversity in pregnancy, family adversity at 0 to 2 years post-partum | logreg | logreg |
| Confounder | Marital status | Maternal | 469 (6.9%) | 1,601 (11.0%) | Paternal social class,  ACE parental separation, Family adversity in pregnancy, family adversity at 0 to 2 years post-partum | Paternal social class, crowding index, ACE physical abuse, ACE emotional abuse, ACE parental substance use, ACE parental violence, ACE parental offence, ACE parental mental health, ACE parental separation, family adversity in pregnancy, family adversity at 0 to 2 years post-partum | logreg | logreg |
| Confounder | Partner alcohol use during pregnancy | Paternal | 1,667 (24.7%) | 4,964 (34.0%) | Weekly alcohol consumption during pregnancy | Crowding index, maternal change in alcohol use during pregnancy, weekly alcohol consumption during pregnancy, maternal self-harm | polyreg | polyreg |
| Exposure | Prenatal alcohol exposure | Maternal | 499 (7.4%) | 1,748 (12.0%) | Financial difficulty during pregnancy, maternal self-harm, weekly alcohol consumption during pregnancy, change in alcohol consumption during pregnancy, alcohol binges in the last month | Financial difficulty during pregnancy, maternal self-harm, weekly alcohol consumption during pregnancy, change in alcohol consumption during pregnancy, alcohol binges in the last month | polyreg | polyreg |
| Exposure | Prenatal binge drinking exposure | Maternal | 485 (7.2%) | 1,733 (11.9%) | Alcohol binges in the last month, weekly alcohol consumption during pregnancy | Alcohol binges in the last month, weekly alcohol consumption during pregnancy | logreg | logreg |
| Outcome | MRB: Alcohol use by 16 years | Child | 2,226 (33.0%) | 10,077 (69.0%) | Clinic reported daily smoking, clinical reported alcohol use | Crowding index, sensation seeking score A, clinic reported daily smoking, clinic reported alcohol use | pmm | logreg |
| Outcome | MRB: Drug use by 16 years | Child | 1,918 (28.4%) | 9,769 (66.9%) | Clinic reported daily smoking | Sensation seeking score A, ACE bullying, ACE parental mental health, ACE emotional abuse, ACE physical abuse, Sex of child, SDQ conduct problems, crowding index, clinic reported daily smoking, clinic reported alcohol use | pmm | logreg |
| Outcome | MRB: Cannabis use by 16 years | Child | 1,713 (25.4%) | 9,564 (65.4%) | Clinic reported daily smoking, clinical reported alcohol use | Clinic reported alcohol use, clinic reported daily smoking | pmm | logreg |
| Outcome | MRB: Smoking by 16 years | Child | 1,716 (25.4%) | 9,567 (65.5%) | Clinic reported daily smoking, clinical reported alcohol use | Sex of child, crowding index, ACE emotional abuse, ACE parental mental health, ACE parental offence, ACE physical abuse, sensation seeking score B, clinic reported daily smoking, clinic reported alcohol use | pmm | logreg |
| Outcome | MRB: Crime or antisocial behaviour by 16 years | Child | 1,413 (20.9%) | 9,264 (63.4%) | Sensation seeking score A, Clinic reported daily smoking, clinical reported alcohol use | ACE emotional abuse, ACE sexual abuse, ACE parental substance use, ACE physical abuse, sensation seeking score A, clinic reported alcohol use, clinic reported daily smoking, Age of child at clinic | pmm | logreg |
| Outcome | MRB: Unprotected sex by 16 years | Child | 1,593 (23.6%) | 9,444 (64.7%) | Clinic reported alcohol use, sensation seeking score A, ACE sexual abuse | Clinic reported alcohol use, sensation seeking score A, ACE sexual abuse | pmm | logreg |
| Outcome | MRB: Underage sexual intercourse | Child | 1,590 (23.5%) | 9441 (64.7%) | Clinic reported alcohol use | Paternal social class, clinic reported alcohol use, ACE emotional abuse, ACE neighbourhood, sensation seeking score A | pmm | logreg |
| Auxiliary variable | Paternal social class | Paternal | 1,029 (15.2%) | 3703 (25.4%) | Crowding index | Financial difficulties during pregnancy, Weekly alcohol consumption during pregnancy, crowding index, Change in alcohol consumption during pregnancy, Neighbourhood index | logreg | logreg |
| Auxiliary variable | Crowding index | Maternal | 586 (8.7%) | 1,883 (12.9%) | Paternal social class, change in alcohol consumption during pregnancy, family adversity during pregnancy, family adversity at 0-2 years post-partum | Paternal social class, change in alcohol consumption during pregnancy, SDQ depression, SDQ hyperactivity, SDQ emotional symptoms, SDQ peer problems, SDQ conduct problems, SDQ total problems, ACE physical abuse, ACE sexual abuse, ACE emotional abuse, ACE bullying, ACE parental violence, ACE parental substance use, ACE parental mental health, ACE parental offence, ACE parental separation, ACE neighbourhood, family adversity during pregnancy | polyreg | polyreg |
| Auxiliary variable | Crown crisp anxiety | Maternal | 975 (14.4%) | 2,901 (19.9%) | Crown crisp depression, ACE parental mental health, family adversity during pregnancy, family adversity at 0-2 years post-partum | Physical activity, Paternal social class, crown crisp depression, financial difficulties during pregnancy, ACE parental mental health, family adversity during pregnancy | polyreg | polyreg |
| Auxiliary variable | Crown crisp depression | Maternal | 1,004 (14.9%) | 2,975 (20.4%) | Crown crisp anxiety, ACE parental mental health, family adversity during pregnancy, family adversity at 0-2 years post-partum | Crown crisp anxiety, financial difficulties during pregnancy, SDQ total problems, ACE parental mental health, family adversity during pregnancy | polyreg | polyreg |
| Auxiliary variable | Financial difficulties during pregnancy | Maternal | 927 (13.7%) | 2,778 (19.0%) | Family adversity during pregnancy, family adversity at 0-2 years post-partum | Maternal self-harm, Change in alcohol consumption during pregnancy, crown crisp anxiety, crown crisp depression, ACE parental mental health, family adversity during pregnancy | polyreg | polyreg |
| Auxiliary variable | Weekly alcohol consumption during pregnancy | Maternal | 3,147 (46.6%) | 7,760 (53.1%) | Alcohol binges in the last month | Change in alcohol consumption during pregnancy, Alcohol binges in the last month | logreg | logreg |
| Auxiliary variable | BMI | Maternal | 925 (13.7%) | 3,142 (21.5%) | No auxiliary variables identified | Neighbourhood index | polyreg | polyreg |
| Auxiliary variable | Neighbourhood index | Maternal | 766 (11.3%) | 2,089 (14.3%) | Correlated variables related to missingness are in the substantive model | Correlated variables related to missingness are in the substantive model | polyreg | polyreg |
| Auxiliary variable | SDQ depression | Child | 1,017 (15.1%) | 6,587 (45.1%) | SDQ emotional problems, SDQ hyperactivity, SDQ peer problems, SDQ conduct problems | SDQ emotional problems, SDQ hyperactivity, SDQ peer problems, SDQ conduct problems | polyreg | polyreg |
| Auxiliary variable | SDQ hyperactivity | Child | 1,067 (15.8%) | 6,590 (45.1%) | SDQ emotional problems, SDQ depression, SDQ peer problems, SDQ conduct problems | SDQ emotional problems, SDQ depression, SDQ peer problems, SDQ conduct problems | polyreg | polyreg |
| Auxiliary variable | SDQ emotional problems | Child | 1,077 (16.0%) | 6,606 (45.2%) | SDQ hyperactivity, SDQ depression, SDQ peer problems, SDQ conduct problems | SDQ hyperactivity, SDQ depression, SDQ peer problems, SDQ conduct problems | polyreg | polyreg |
| Auxiliary variable | SDQ peer problems | Child | 1,076 (15.9%) | 6,600 (45.2%) | SDQ emotional problems, SDQ depression, SDQ hyperactivity, SDQ conduct problems | SDQ emotional problems, SDQ depression, SDQ hyperactivity, SDQ conduct problems | polyreg | polyreg |
| Auxiliary variable | SDQ conduct problems | Child | 1,069 (15.8%) | 6,590 (45.1%) | SDQ emotional problems, SDQ hyperactivity, SDQ peer problems, SDQ depression | SDQ emotional problems, SDQ hyperactivity, SDQ peer problems, SDQ depression | polyreg | polyreg |
| Auxiliary variable | SDQ total problems | Child | 1,085 (16.1%) | 6,619 (45.3%) | SDQ depression, SDQ emotional problems, SDQ hyperactivity, SDQ peer problems, SDQ conduct problems | SDQ depression, SDQ emotional problems, SDQ hyperactivity, SDQ peer problems, SDQ conduct problems | polyreg | polyreg |
| Auxiliary variable | ACE physical abuse | Child | 1,982 (29.3%) | 8,200 (56.2%) | ACE emotional abuse | Sensation seeking score B, ACE emotional abuse, age of child at clinic, crowding index | logreg | logreg |
| Auxiliary variable | ACE sexual abuse | Child | 873 (12.9%) | 5,542 (38.0%) | No auxiliary variables identified | No auxiliary variables identified | logreg | logreg |
| Auxiliary variable | ACE emotional abuse | Child | 1,818 (26.9%) | 7,727 (52.9%) | ACE physical abuse, ACE parental violence, ACE parental mental health, ACE parental separation | ACE physical abuse, ACE parental violence, ACE parental mental health, ACE parental separation | logreg | logreg |
| Auxiliary variable | ACE bullying | Child | 1,129 (16.7%) | 7,566 (51.8%) | No auxiliary variables identified | ACE parental substance use, SDQ emotional symptoms | logreg | logreg |
| Auxiliary variable | ACE parental violence | Child | 2,000 (29.6%) | 8,234 (56.4%) | ACE parental separation, ACE emotional abuse, family adversity at 0-2 years post-partum | Paternal social class, SDQ total problems, sensation seeking score A, ACE parental separation, ACE emotional abuse, SDQ conduct problems | logreg | logreg |
| Auxiliary variable | ACE parental substance use | Child | 1,627 (24.1%) | 7,280 (49.9%) | Correlated variables related to missingness are in the substantive model | Correlated variables related to missingness are in the substantive model | logreg | logreg |
| Auxiliary variable | ACE parental mental health | Child | 1,456 (21.6%) | 7,271 (49.8%) | Crown crisp anxiety, crown crisp depression, ACE emotional abuse, family adversity at 0-2 years post-partum | Crown crisp anxiety, crown crisp depression, ACE emotional abuse | logreg | logreg |
| Auxiliary variable | ACE parental offence | Child | 1,460 (21.6%) | 6,999 (47.9%) | No auxiliary variables identified | No auxiliary variables identified | logreg | logreg |
| Auxiliary variable | ACE parental separation | Child | 1,885 (27.9%) | 8,044 (55.1%) | ACE emotional abuse, ACE parental violence | Age of child at clinic, ACE parental substance use, ACE emotional abuse, ACE parental violence | logreg | logreg |
| Auxiliary variable | ACE neighbourhood | Child | 504 (7.5%) | 5,850 (40.1%) | No auxillary variables identified | No auxiliary variables identified | logreg | logreg |
| Auxiliary variable | Antisocial activities | Child | 1,541 (22.8%) | 7,514 (51.5%) | No auxillary variables identified | Age of child at clinic | logreg | logreg |
| Auxiliary variable | Sensation seeking score A | Child | 1,304 (19.3%) | 7,650 (52.4%) | Sensation seeking score B | Sensation seeking score B, Sex of child | polyreg | polyreg |
| Auxiliary variable | Sensation seeking score B | Child | 1,314 (19.5%) | 7,660 (52.5%) | Sensation seeking score A | Sensation seeking score A, ACE neighbourhood | polyreg | polyreg |
| Auxiliary variable | Change in alcohol consumption during pregnancy | Maternal | 893 (13.2%) | 2,913 (19.9%) | No auxiliary variables identified | Weekly alcohol consumption during pregnancy | polyreg | polyreg |
| Auxiliary variable | Alcohol binges during the last month | Maternal | 2,315 (34.2%) | 5,932 (40.6%) | Weekly alcohol consumption during pregnancy | Weekly alcohol consumption during pregnancy | polyreg | polyreg |
| Auxiliary variable | Clinic reported alcohol use | Child | 2,237 (33.1%) | 10,027 (68.7%) | Clinic reported daily smoking | ACE emotional abuse, SDQ depression, SDQ peer problems, SDQ hyperactivity, SDQ hyperactivity, SDQ conduct problems | logreg | logreg |
| Auxiliary variable | Sex of child | Child | 0 (0.0%) | 0 (0.0%) | Sensation seeking score A, child reported self-harm | Sensation seeking score A | logreg | logreg |
| Auxiliary variable | Maternal self-harm | Maternal | 838 (12.4%) | 2,579 (17.7%) | No auxiliary variables identified | No auxiliary variables identified | polyreg | polyreg |
| Auxiliary variable | Physical activity per week | Maternal | 1,105 (16.4%) | 3,333 (22.8%) | Alcohol binges during the last month, Change in alcohol consumption during pregnancy, family adversity during pregnancy | Alcohol binges during the last month, Change in alcohol consumption during pregnancy | polyreg | polyreg |
| Auxiliary variable | Age of child at clinic | Child | 1,364 (20.2%) | 9120 (62.5%) | ACE bullying, sensation seeking score A, sensation seeking score B | SDQ depression, SDQ hyperactivity, SDQ emotional symptoms, SDQ peer problems, SDQ conduct problems, ACE bullying, ACE parental violence, ACE parental offence, ACE parental separation, sensation seeking score A, sensation seeking score B | polyreg | polyreg |
| Auxiliary variable | Clinic reported daily smoking | Child | 4,593 (68.0%) | 12,444 (85.2%) | Clinic reported alcohol use | Clinic reported alcohol use, Sex of child, crowding index, ACE parental violence, ACE parental mental health, sensation seeking score A, sensation seeking score B | logreg | logreg |
| Auxiliary variable | Family adversity during pregnancy | Maternal | 1,195 (17.7%) | 3,922 (26.9%) | Crowding index, crown crisp depression, crown crisp anxiety, family adversity at 0-2 years post-partum, ACE emotional abuse | Crowding index, crown crisp depression, crown crisp anxiety | polyreg | polyreg |
| Auxiliary variable | Family adversity family adversity at 0 to 2 years post-partum | Maternal | 311 (4.6%) | 716 (4.9%) | Crowding index, crown crisp depression, crown crisp anxiety, family adversity during pregnancy, ACE emotional abuse, ACE parental mental health, ACE parental violence | Crowding index, crown crisp depression, crown crisp anxiety, SDQ child total problems, ACE emotional abuse, ACE parental mental health, ACE parental violence, family adversity during pregnancy | polyreg | polyreg |
| Auxiliary variable | Child self-harm | Child | 1,704 (25.2%) | 9,555 (65.4%) | Clinic reported daily smoking | Sex of child, clinic reported daily smoking | logreg | logreg |
| Outcome | MRB: Total score (out of 7) | Child | 4,747 (70.3%) | 12,598 (86.2%) | No auxiliary variables used as passively imputed as the sum of individual MRBs | No auxiliary variables used as passively imputed as the sum of individual MRBs | ~I(alcohol_16 + drugs_16 + cannabis_16 + smoke_16 + crimeasb_16 + sexnocont_16 + underage_16) | Not included |

**Multiple imputation stability for outcomes**

**Supplementary Figure A. Mean MRB score before adjustment and standard deviation across multiple imputation iterations (full imputation)**

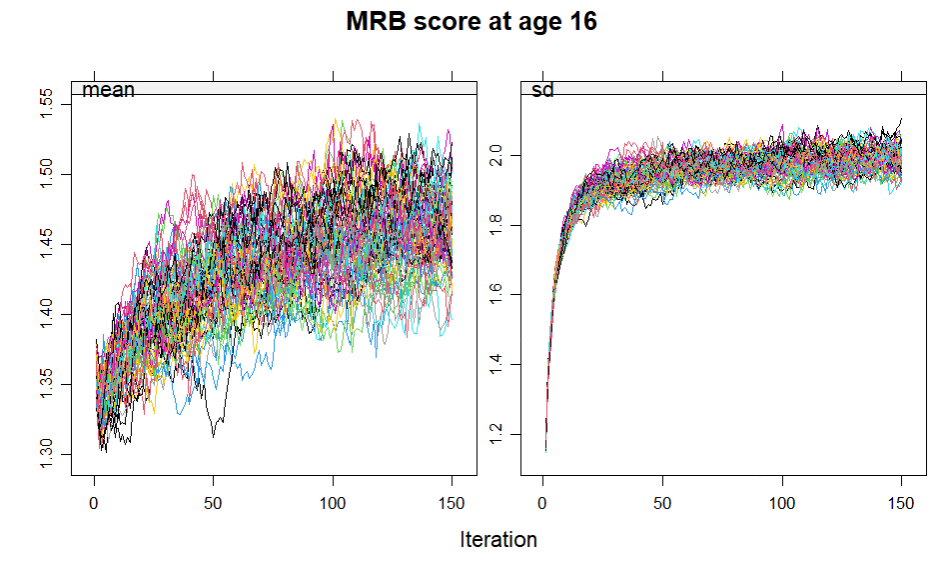

**Supplementary Figure B. Mean value between 1 (No) and 2 (Yes) and standard deviation for hazardous alcohol use at age 16 years (full imputation)**

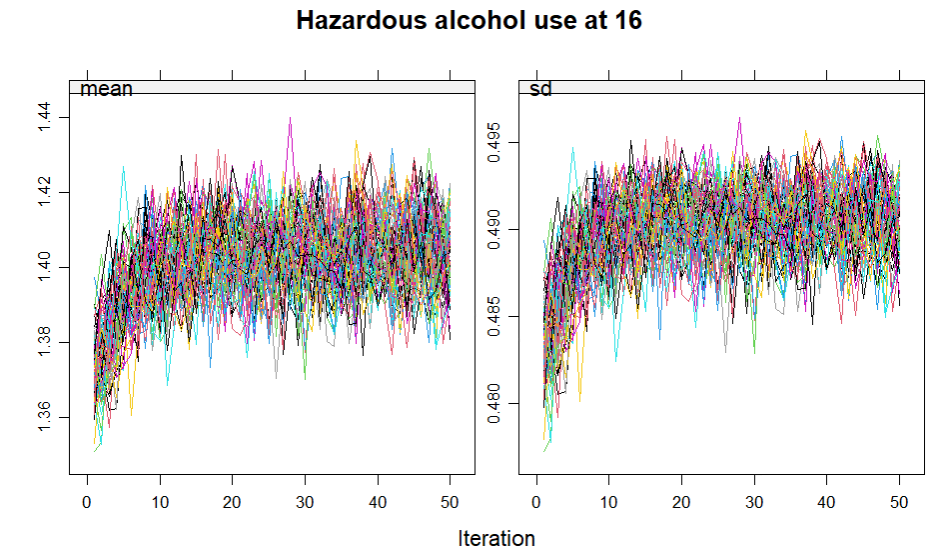

**Supplementary Figure C. Mean value between 1 (No) and 2 (Yes) and standard deviation for regular smoking at age 16 (full imputation)**

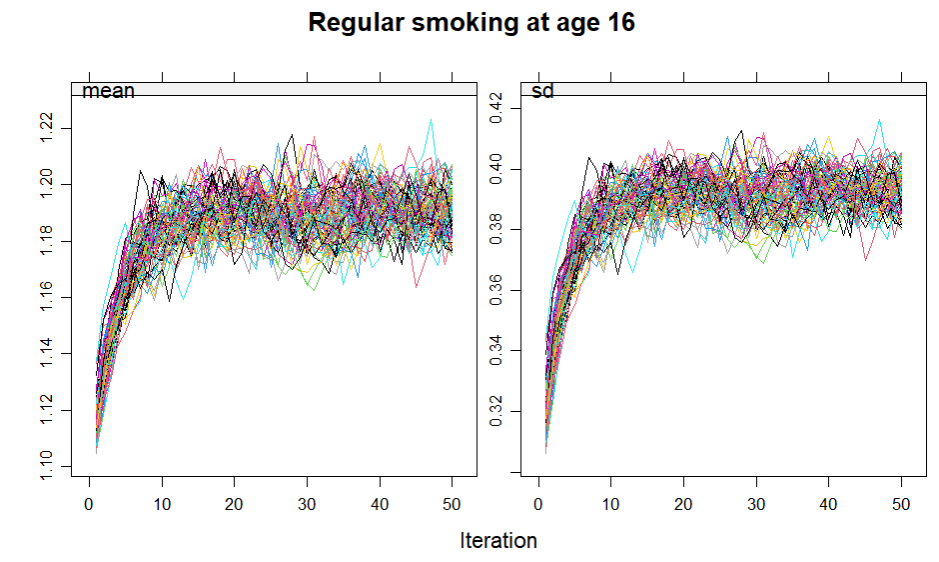

**Supplementary Figure D. Mean value between 1 (No) and 2 (Yes) and standard deviation for drug use at age 16 (full imputation)**

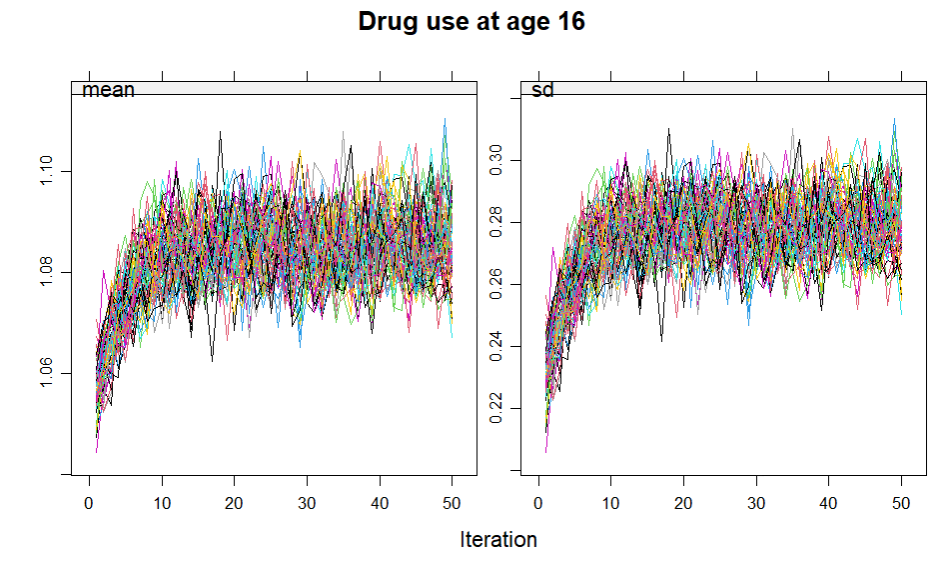

**Supplementary Figure E. Mean value between 1 (No) and 2 (Yes) and standard deviation for regular cannabis use at age 16 (full imputation)**

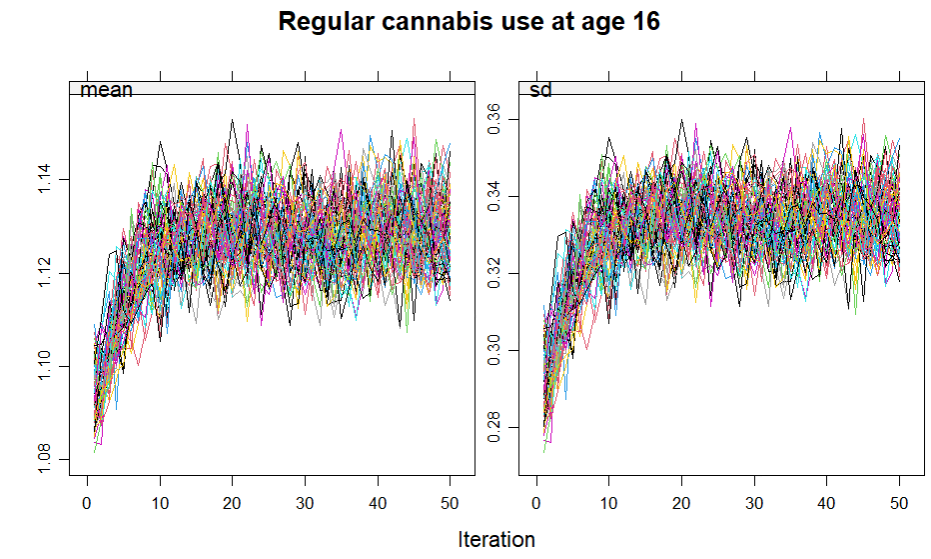

**Supplementary Figure F. Mean value between 1 (No) and 2 (Yes) and standard deviation for crime or antisocial behaviour by age 16 (full imputation)**

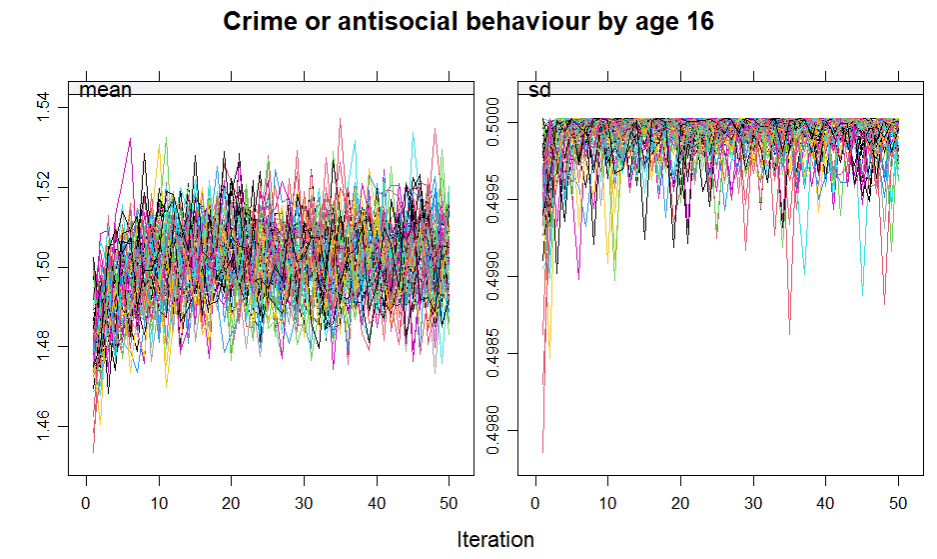

**Supplementary Figure G. Mean value between 1 (No) and 2 (Yes) and standard deviation for unprotected sex by age 16 (full imputation)**

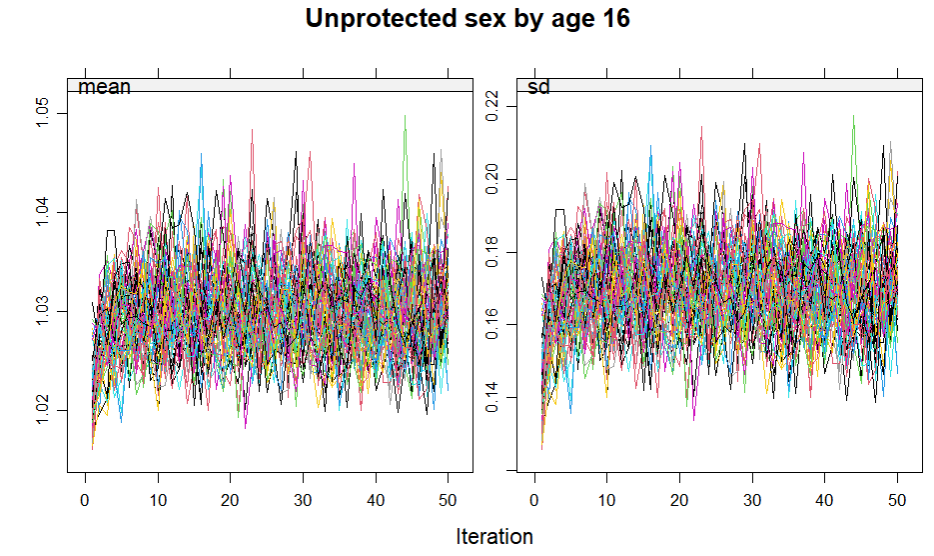

**Supplementary Figure H. Mean value between 1 (No) and 2 (Yes) and standard deviation for underage sex before 16^th^ birthday (full imputation)**

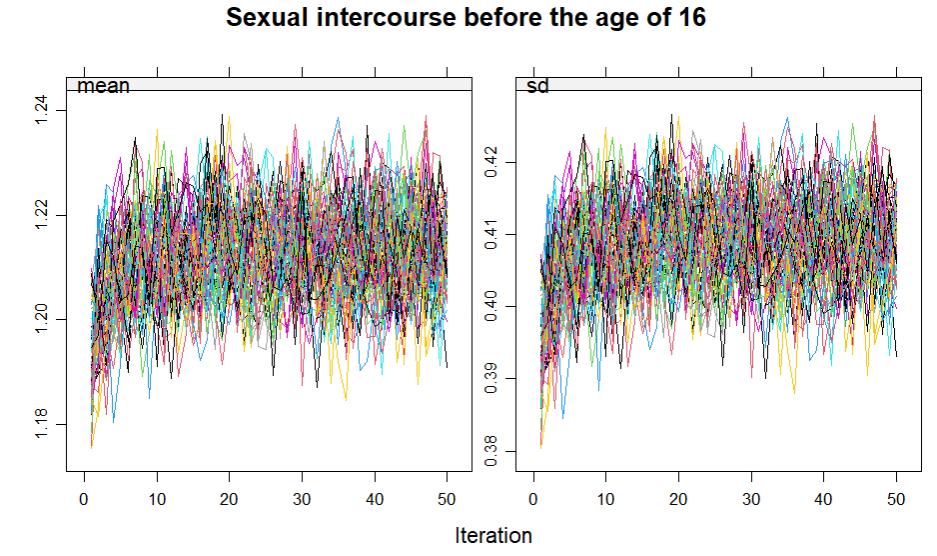

**Supplementary Figure I. Mean MRB score before adjustment and standard deviation across multiple imputation iterations (partial imputation)**
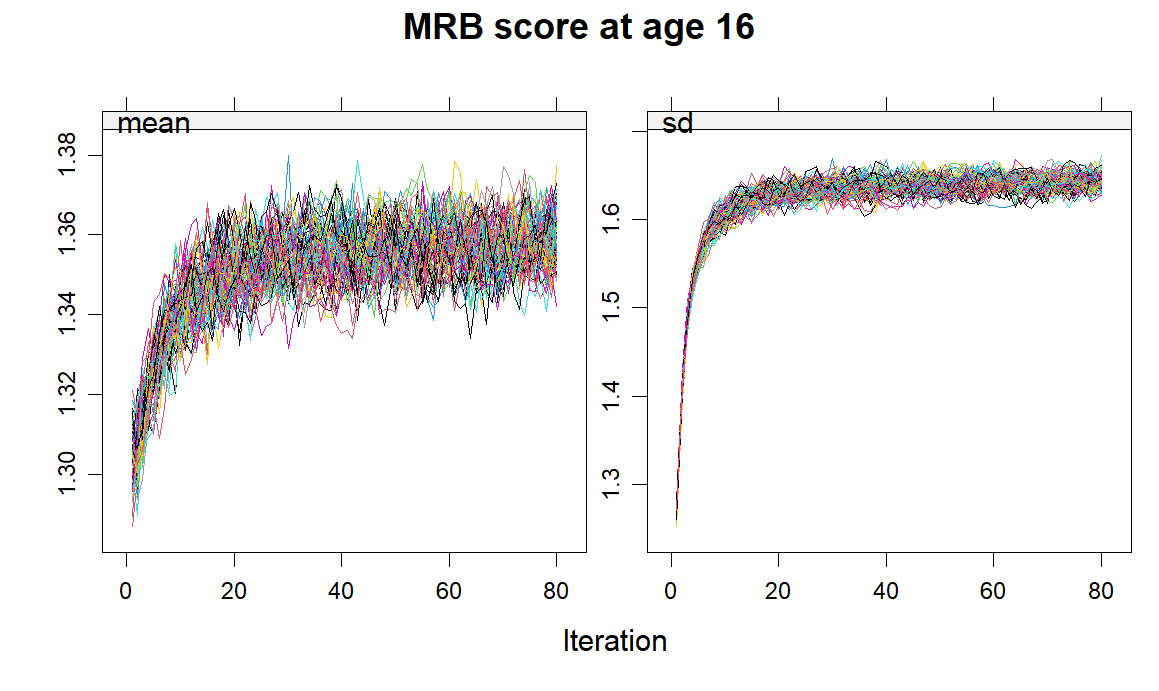

**Supplementary Figure J. Mean value between 1 (No) and 2 (Yes) and standard deviation for hazardous alcohol use at age 16 years (partial imputation)**

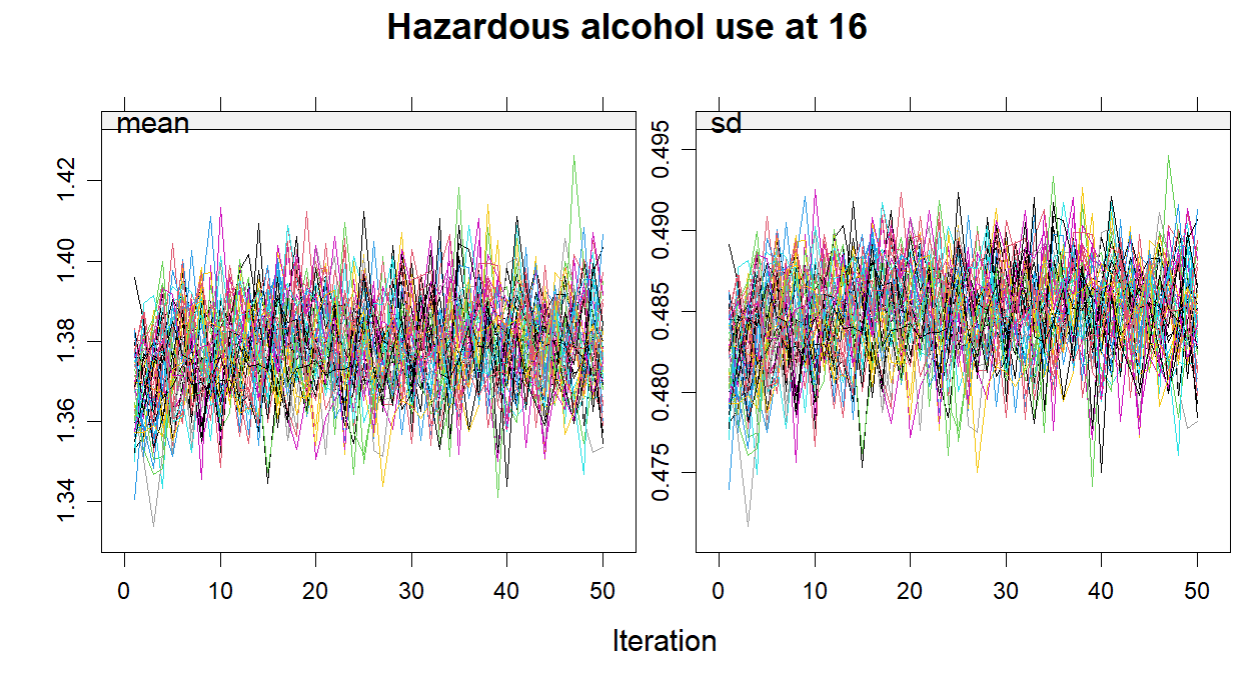

**Supplementary Figure K. Mean value between 1 (No) and 2 (Yes) and standard deviation for regular smoking at age 16 (partial imputation)**

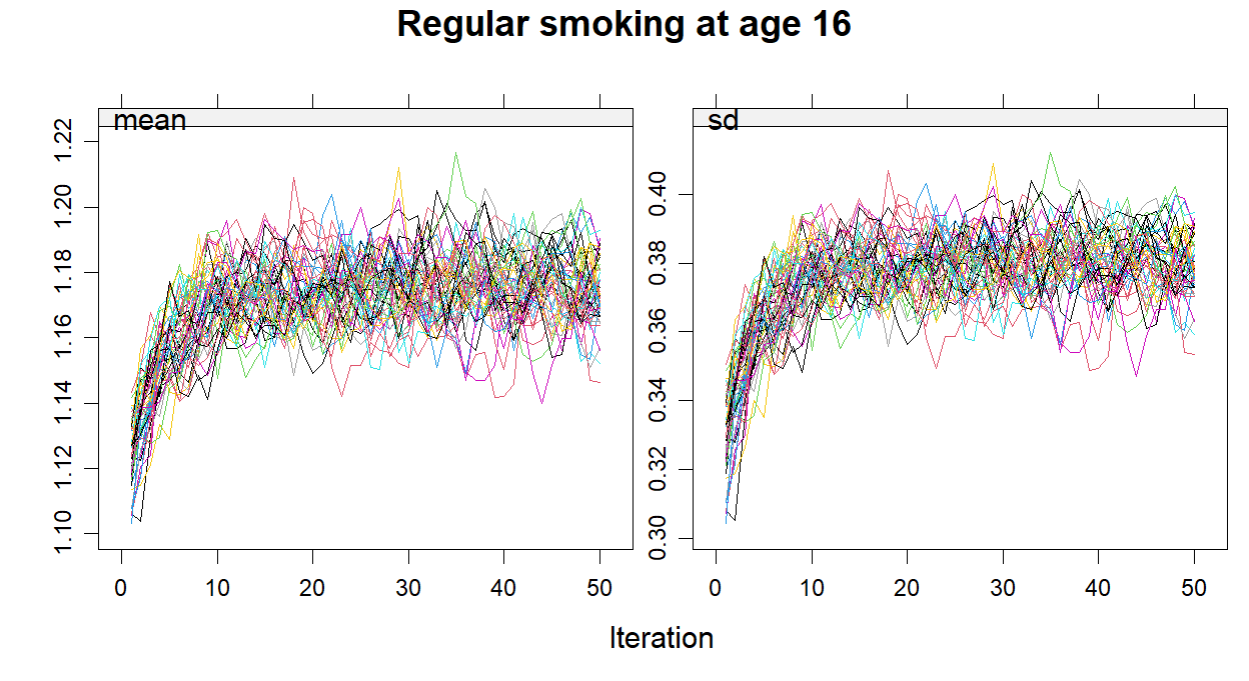

**Supplementary Figure L. Mean value between 1 (No) and 2 (Yes) and standard deviation for drug use at age 16 (partial imputation)**

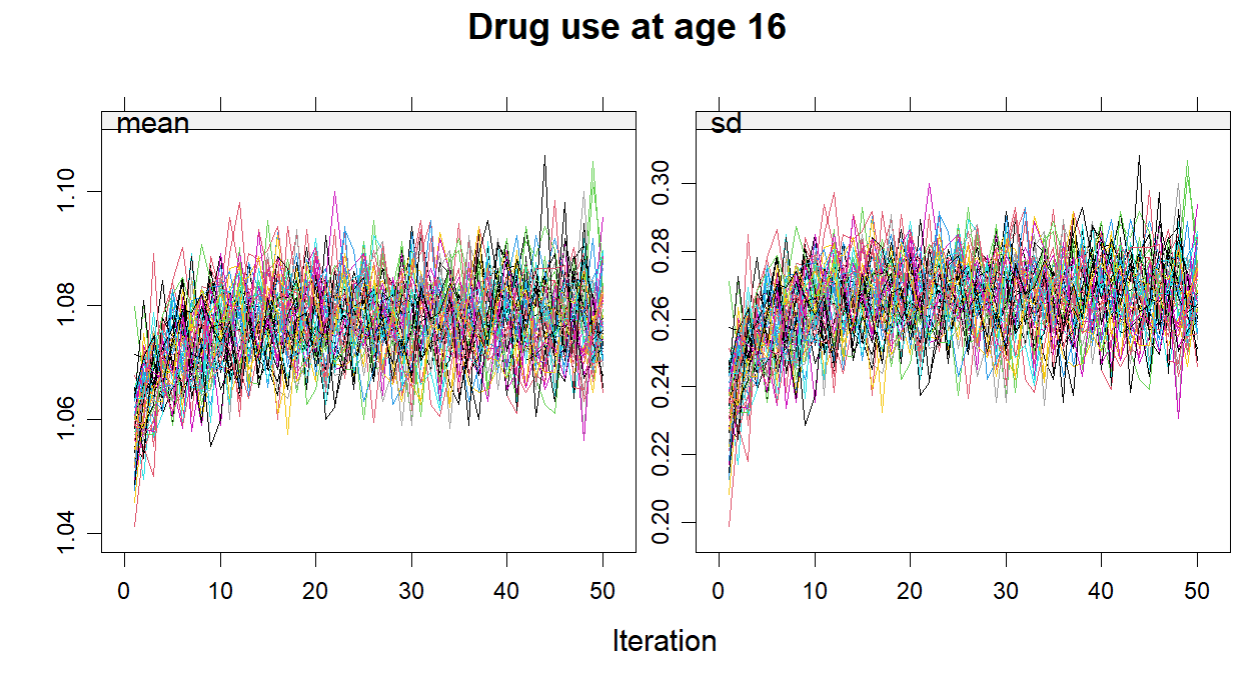

**Supplementary Figure M. Mean value between 1 (No) and 2 (Yes) and standard deviation for regular cannabis use at age 16 (partial imputation)**

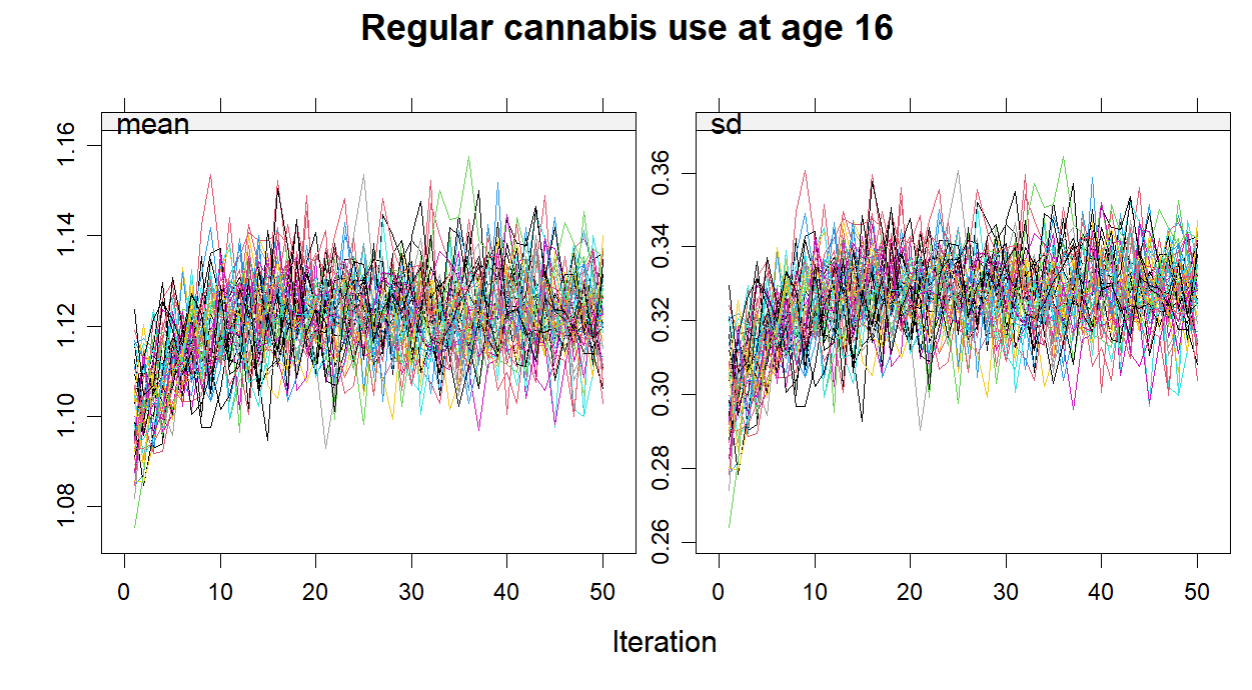

**Supplementary Figure N. Mean value between 1 (No) and 2 (Yes) and standard deviation for crime or antisocial behaviour by age 16 (partial imputation)**

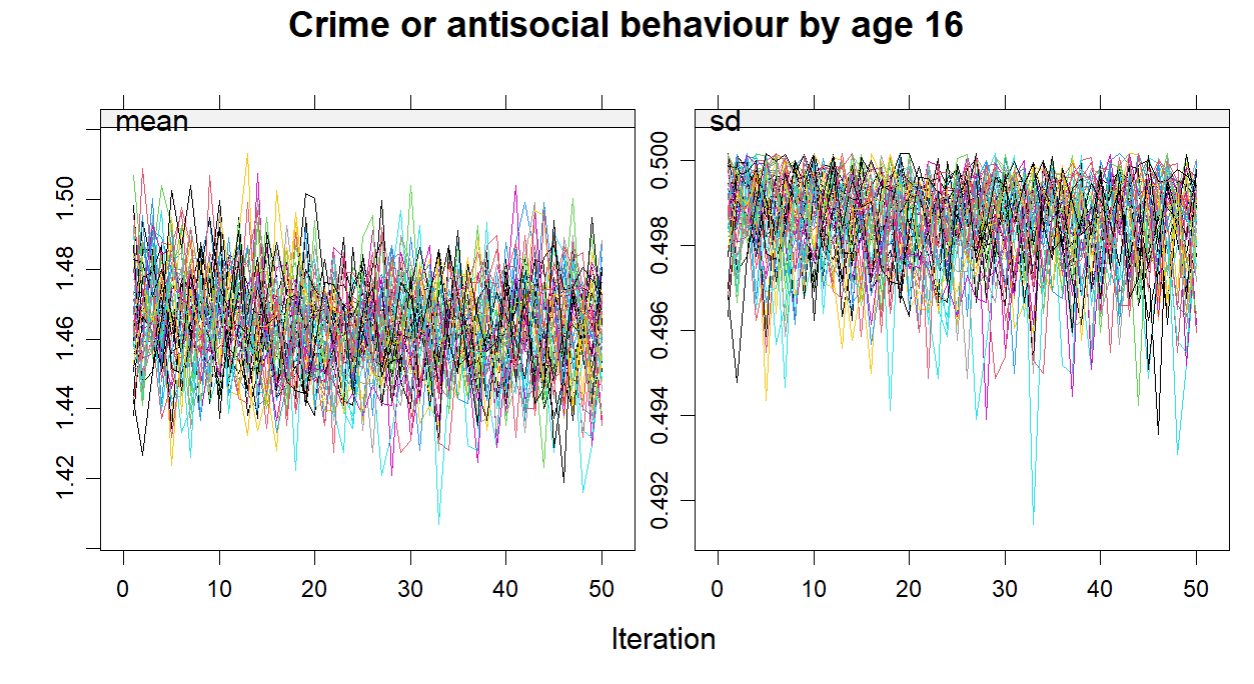

**Supplementary Figure O. Mean value between 1 (No) and 2 (Yes) and standard deviation for unprotected sex by age 16 (partial imputation)**

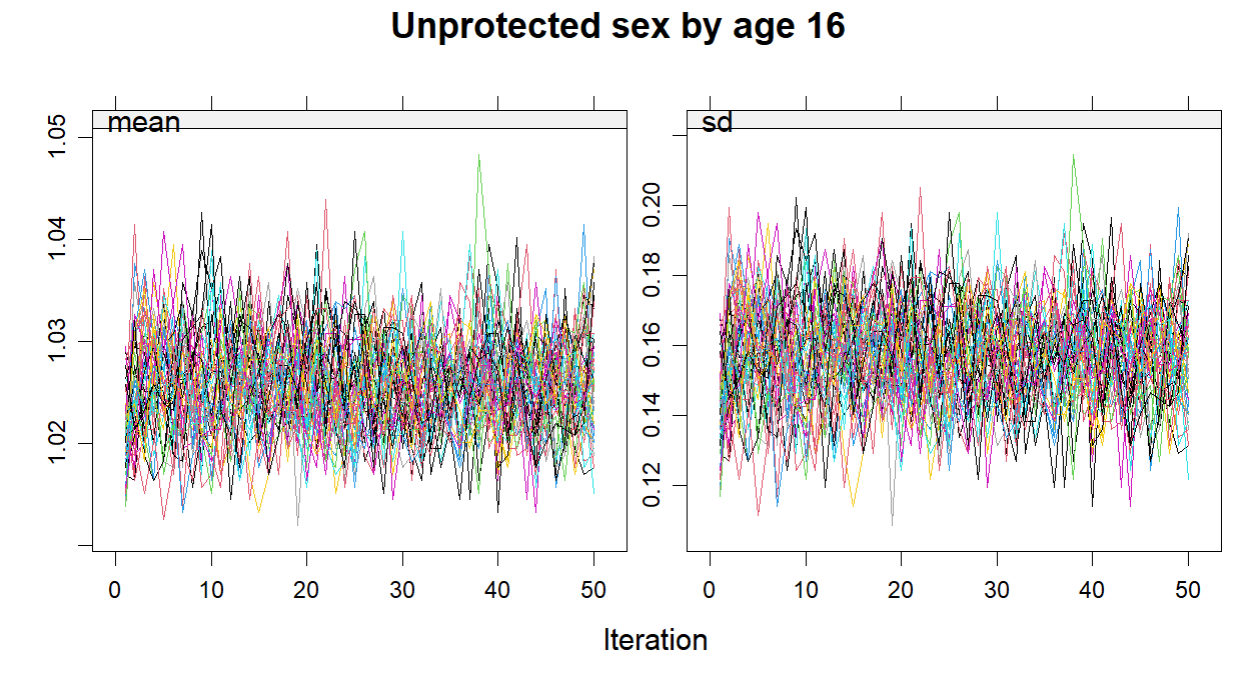

**Supplementary Figure P. Mean value between 1 (No) and 2 (Yes) and standard deviation for underage sex before 16^th^ birthday (partial imputation)**

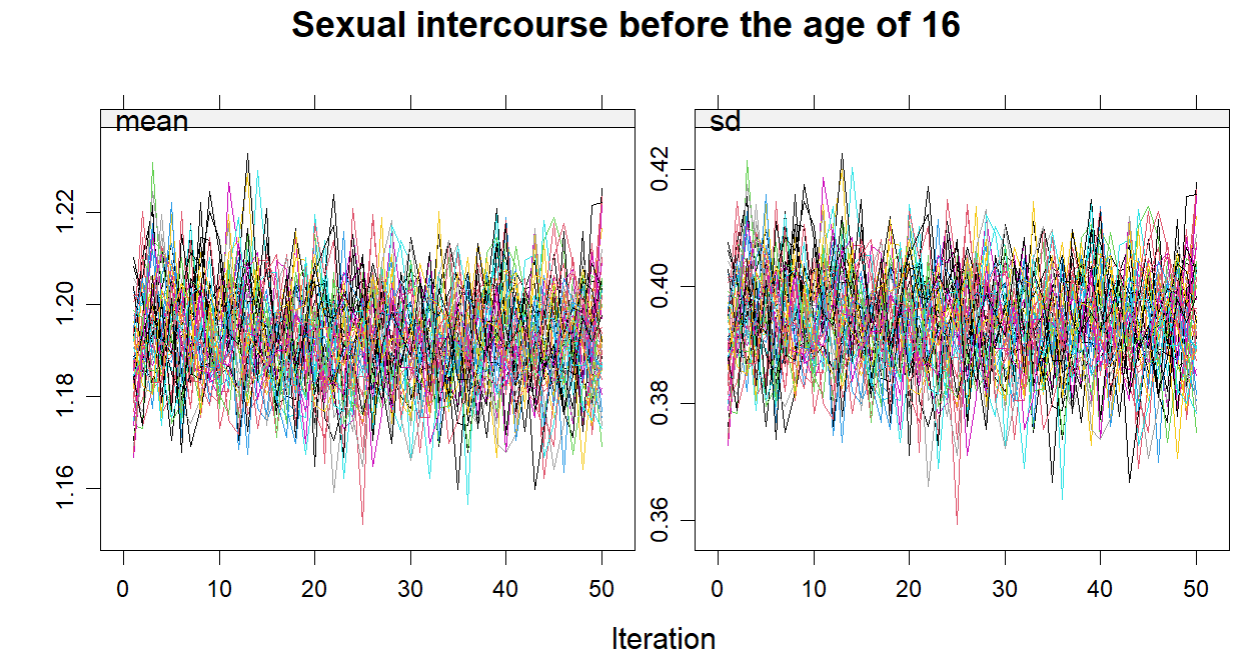
